# Long-term outcomes of COVID-19 infection in children and young people: a systematic review and meta-analysis

**DOI:** 10.1101/2023.04.04.23288110

**Authors:** Helen Twohig, Ram Bajpai, Nadia Corp, Alice Faux-Nightingale, Christian Mallen, Toni Robinson, Glenys Somayajula, Danielle Van der Windt, Victoria Welsh, Claire Burton

## Abstract

**Background:** Children and young people (CYP) may experience prolonged symptoms following COVID-19, commonly termed ‘Long-COVID’. The nature of this in CYP is unclear, as are the sequalae of acute COVID-19. We aimed to systematically synthesise evidence of the long-term outcomes of COVID-19 in CYP.

**Methods:** 13 databases were searched until January 2022. Inclusion criteria: Observational studies reporting outcomes occurring four-weeks or more after COVID-19 in children <18 years old. Exclusion criteria: Outcomes of Paediatric Inflammatory Multisystem Syndrome. Title, abstract and full text screening were conducted independently by two reviewers. Data extraction and risk of bias assessment was by one reviewer with independent verification. Critical appraisal tools appropriate for study type were employed. Results were narratively synthesised with meta-analysis to generate summary estimates of risk of prolonged symptoms in CYP.

**Findings:** 94 studies were included. 66 recruited from hospital settings, 8 recruited solely from community settings. >100 symptoms were reported, the most common being fatigue, headache and cognitive symptoms. Summary estimates of risk of prolonged symptoms were higher for hospital samples (31.2%, 95% CI 20.3% to 43.2%) than for community samples (4.6%, 95% CI 3.4% to 5.8). Sequalae including stroke, type-1 diabetes, Guillan-Barre syndrome, and persistent radiological or blood test abnormalities have been reported in CYP following COVID-19. Most studies reporting these are case reports / case series and quality of evidence is low.

**Interpretation:** Prolonged symptoms following COVID-19 in children are variable and multi-system. Rates in community samples are lower than hospital. There is limited data on other sequalae in CYP. Heterogeneity in diagnosis of COVID-19, symptom classification, assessment method and duration of follow-up made synthesis less secure.

**Funding:** HT, CB and GS have National Institute for Health and Care Research fellowships. RB, CM and VW are supported by the NIHR West Midlands Applied Research Collaboration. CM Is supported by the NIHR School for Primary Care Research

**Research in context panel:** *Evidence before this study:* At the time of writing and to the best of our knowledge, the protocol for this systematic review was a novel endeavour to summarise the longer-term effects of COVID-19 in children and young people (CYP). At least three systematic reviews have since been published, summarising the symptom profile and prevalence of Long-COVID in CYP, but prevalence estimates vary widely and the evidence base remains uncertain. In addition, there is very limited information on other sequalae of COVID-19 in this population group. We searched thirteen electronic databases (MEDLINE, EMBASE, AMED, HMIC, CINAHLPlus, PsycINFO, Web of Science (Science Citation and Social Science Citation indicies), ASSIA, WHO COVID-19: Global literature on coronavirus disease, Cochrane COVID-19 study register, ProQuest Coronavirus research database, NDLTD and OpenGrey) up to January 2022 for any empirical study including search terms pertaining to longer term symptoms of COVID-19 in CYP (<18 years old). The quality of the studies was mixed. Results were analysed narratively for each objective, and random effects meta-analyses conducted to estimate risk of prolonged symptoms in CYP who have had COVID-19.

*Added value of this study:* This review adds to the evidence of the heterogeneity of prolonged symptoms following COVID-19 in CYP but importantly, stratifies risk of this by recruitment setting. We also synthesise evidence on broader sequalae of the acute infection in this CYP and longer-term effects in CYP with pre-existing conditions, which have not been considered in previous reviews. We purposefully included case studies and case series, to capture emerging patterns of outcomes, which may well be important in a novel condition with a rapidly increasing volume of publications. To our knowledge, this systematic review and meta-analysis is the most comprehensive to date.

*Implications of all the available evidence:* This review adds to the evidence that a substantial proportion of CYP do experience effects of COVID-19 that last longer than four-weeks, with the most frequently reported prolonged symptoms being fatigue, headache and cognitive symptoms. The proportion of CYP developing prolonged symptoms in children recruited from community setting was low, although this may translate to a large number of affected CYP at population level. There is a paucity of controlled studies and this limits confidence that prolonged symptoms are attributable to COVID-19. Sequalae including stroke, type-1 diabetes, Guillan-Barre syndrome, and persistent radiological or blood test abnormalities have been reported in CYP following COVID-19 but most studies reporting these are case reports / case series and quality of evidence is low. To develop treatment plans and interventions for affected CYP, further studies are needed to better characterise this condition and understand its impact on the lives of CYP and their families and communities. These should ideally recruit from community settings, include population-based control groups and consider using standardised definitions and outcome measures where possible.

## Introduction

Acute SARS-CoV-2 infection (COVID-19) in children and young people (CYP) typically presents as a mild illness resulting in fewer hospitalisations, complications, or deaths than in adults1. Fever and cough are the most common symptoms, with other symptoms including rhinorrhoea, sore throat, headache, fatigue/myalgia and gastrointestinal symptoms occurring in less than 10-20%.^2^

It became clear early in the pandemic that some people experience prolonged symptoms following COVID-19^3,4^. Definitions of this condition vary within the literature^5,6^ although the patient derived term^7^, Long-COVID, is well-established. The National Institute for Health and Care Excellence (NICE) use the clinical case definitions of ongoing symptomatic (or post-acute) Covid-19 (5-12 weeks after onset) and post-Covid-19 syndrome (symptoms lasting 12 weeks or more)8 but acknowledge that ‘Long-COVID’ is commonly used to encompass both. Research into Long-COVID has predominantly focused on adults and there is limited literature on the condition in CYP^9,10^.

Three reviews have reported clinical presentation and prevalence of Long-COVID in CYP^11,12,13^, though their findings range from no difference in persistent symptoms between cases and control groups^11^, to a higher risk of some persistent symptoms in cases compared to controls^12^, and a narrative synthesis finding that prevalence estimates and symptom burden varied widely13. Reported ‘prevalence’ in these reviews refers to risk of prolonged symptoms in children who had COVID-19, rather than population prevalence.

Studies in adult post-COVID populations have demonstrated risks of complications including cardiovascular events, strokes and venous thrombosis^14-17^ with risks of more severe COVID-19 and complications in people with underlying chronic conditions.^18^ These areas have not been well studied in CYP.

Identifying the most common symptoms and sequalae that CYP experience following COVID-19 and estimating their frequency will help clinicians to recognise these complications and implement personalised management. Such information is also necessary to inform development and commissioning of appropriate services. We aimed to synthesise knowledge of longer-term effects of SARS-CoV-2 infection on CYP. Specific objectives were to identify; 1) patterns of symptoms lasting longer than four-weeks 2) risk of developing prolonged symptoms, 3) other sequalae of the infection (including clinical effects and persistent radiological and pathological findings) and 4) risk of longer-term effects in children with pre-existing long-term conditions.

## Methods

This systematic review is registered with PROSPERO (<u>Registration number CRD42020226624</u>) and is reported in accordance with Preferred Reporting Items for Systematic Review and Meta-analyses (PRISMA) 2020 guidelines^19^ (Table S1). We included studies published between December 2019 and January 2022 that reported outcomes at four-weeks or beyond in children aged 0 to 18 years old who had had COVID-19 (positive antigen test, or clinical diagnosis of COVID-19). Pre-prints were included if they were published in a peer-reviewed journal before November 1^st^, 2022.

Studies were excluded where duration of symptoms was unclear, or less than four-weeks, although studies reporting development of new chronic morbidities secondary to COVID-19 infection were included despite short follow-up duration (e.g. diabetes mellitus, stroke sequelae). Studies including adults were excluded unless data pertaining to children were separately presented. Studies solely reporting data on children affected by Paediatric Inflammatory Multisystem Syndrome Temporally Associated with SARS-CoV-2 (PIMS-TS) were excluded, given the unique nature of this rare and severe complication. Primary research of any design, any setting and in any language was included. Secondary evidence, conference abstracts and trial protocols were excluded.

We searched 13 electronic databases (MEDLINE, EMBASE, AMED, HMIC, CINAHLPlus, PsycINFO, Web of Science (Science Citation and Social Science Citation indices), ASSIA, WHO COVID-19: Global literature on coronavirus disease, Cochrane COVID-19 study register, ProQuest Coronavirus research database, NDLTD and OpenGrey) and reference lists of existing systematic reviews. Searches utilised text word searching in the title, abstract and keywords, along with database subject headings, combining terms for paediatrics AND COVID-19 AND long-term (Table S2). Search terms were adapted for each database platform.

Searches were run by NC, on 31^st^ December 2021. Results were imported into Endnote X9 (reference management software, Clarivate Analytics, available at https://endnote.com/) where duplicates were removed. Remaining, unique references were uploaded to Covidence systematic review software (Veritas Health Innovation, Melbourne, Australia. Available at www.covidence.org) to manage the screening process.

Title and abstract followed by full text screening were conducted independently by two reviewers (from CB, HT, VW, GS, TR). Discordance was resolved by team discussion (CB, NC, HT, VW, GS, TR). Main reasons for exclusion during the full text stage were recorded.

Data extraction and risk of bias assessment was by one reviewer, independently checked and validated by a second. We extracted the following data: author; year of publication; setting (country of origin, specific details of community, healthcare setting); study type; study aim; funding; age of participants (range, mean, SD); sex (counts and proportions); number of participants; COVID-19 exposure: definition (including symptom description, antigen test status); details of any comparator group; description of any covariates (e.g. demographic information); potential prognostic factors; outcome measure(s): which have been measured, how have they been measured; follow-up time points; measures of association where relevant.

Risk of bias was assessed using the relevant CASP checklist (https://casp-uk.net/casp-tools-checklists/) for cohort studies and the Joanna Briggs Institute critical appraisal tool (https://jbi.global/critical-appraisal-tools) for case studies and case series. Results of the checklists were used to categorise studies as high, moderate, or low risk of bias.

Studies were grouped into those that reported symptoms lasting more than four-weeks (objectives 1 and 2), those reporting sequalae of the acute infection persisting for more than four-weeks (including clinical diagnoses and persisting pathological or radiological abnormalities in asymptomatic children) (objective 3), and those reporting prolonged symptoms in children with pre-existing long-term conditions (objective 4).

A narrative synthesis was carried out for all groups of studies^20^. For objective 1, where it was possible to discern numbers of children affected by a particular symptom, symptoms were grouped into clinical categories and an infographic was developed to present findings in a clinically meaningful way.

Studies which reported the proportion of children with COVID-19 who experienced prolonged symptoms (objective 2) were included in a meta-analysis to generate summary estimates of the risk of prolonged symptoms in CYP who have had COVID-19. Risk was calculated using the numerator as the number of participants with symptoms lasting longer than four-weeks and the denominator as the total number of participants. The analysis was stratified by recruitment setting (hospital or community) and further by duration of prolonged symptoms (beyond four or twelve weeks).

Individual study prevalence (risk of prolonged symptoms) was pooled using inverse variance DerSimonian and Laird method to fit the random effects model, selected due to the methodological variability across studies. Heterogeneity was summarised using the estimate of between study variance (t2), and the proportion of variability in effect estimates due to between study heterogeneity was summarised using I^2^. A 95% prediction interval for the random-effects model was calculated if at least three studies were available in a meta-analysis 21. Where comparator group data were available, we calculated pooled odds ratio (OR) and 95% CIs using the same methodology.

Publication bias was assessed by visual inspection of a funnel plot if ten or more studies were available for a given outcome. Asymmetry in the publication bias was tested by Egger’s method (p<0·1 was considered as an indication of publication bias)^22^. Meta-analysis and subgroup analysis were performed using Stata v17·0 (Stata-Corp, College Station, Texas, USA). A p-value <0·05 was considered statistically significant for overall and subgroup effects.

### Role of the funding source

The study funder had no role in study design, data collection, data analysis, data interpretation, or writing of the report.

## Results

The study selection process is described in Figure 1. 94 studies were included in the review^23-116^. Characteristics of included studies are summarised in Table 1. Details of all included studies are in supplementary data (S2).

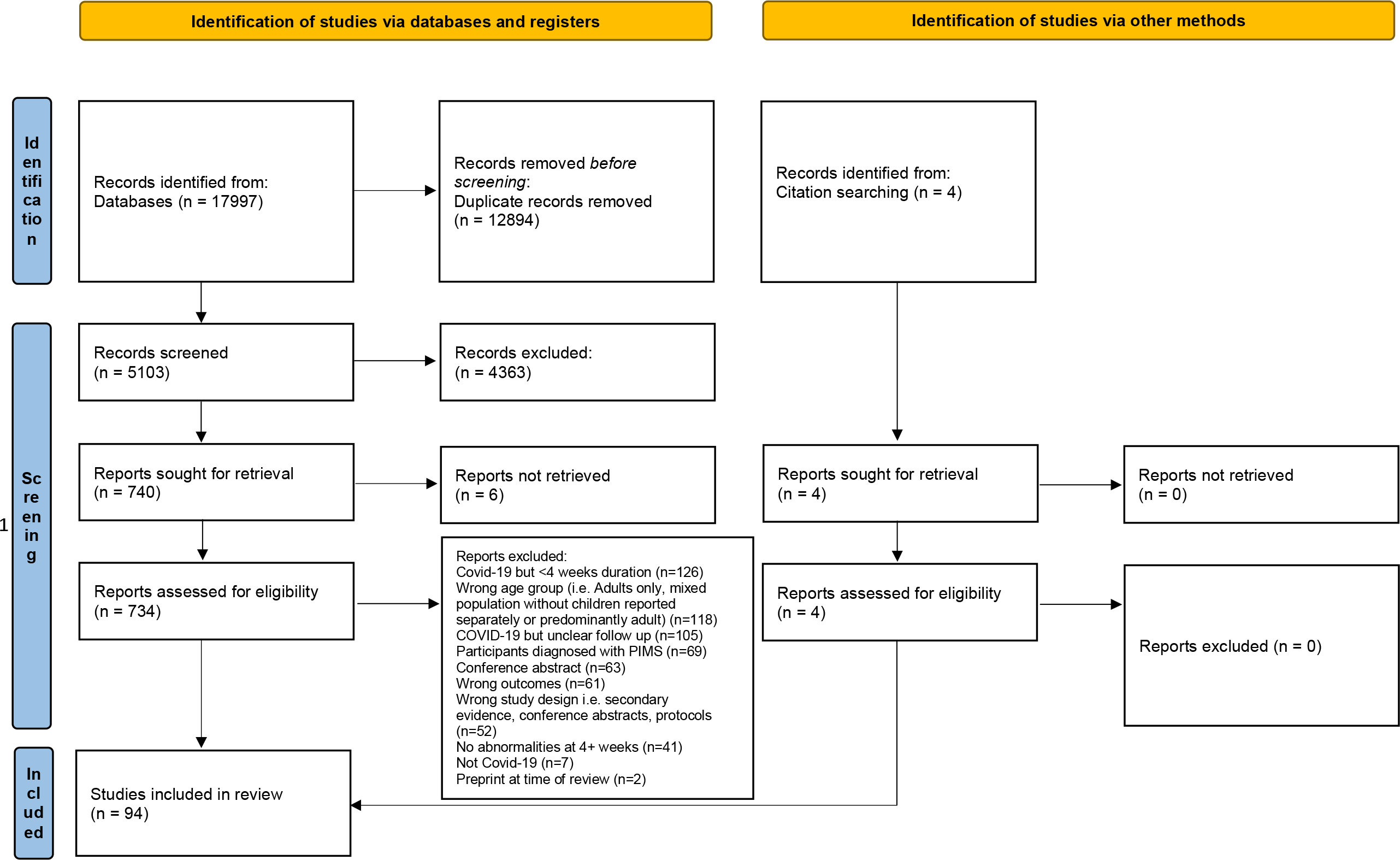
PRISMA 2020 flow diagram for new systematic reviews which included searches of databases, registers and other sources. *From:* Page MJ, McKenzie JE, Bossuyt PM, Boutron I, Hoffmann TC, Mulrow CD, et al. The PRISMA 2020 statement: an updated guideline for reporting systematic reviews. BMJ 2021;372:n71. doi: 10.1136/bmj.n71. For more information, visit: http://www.prisma-statement.org/

**Table 1:**
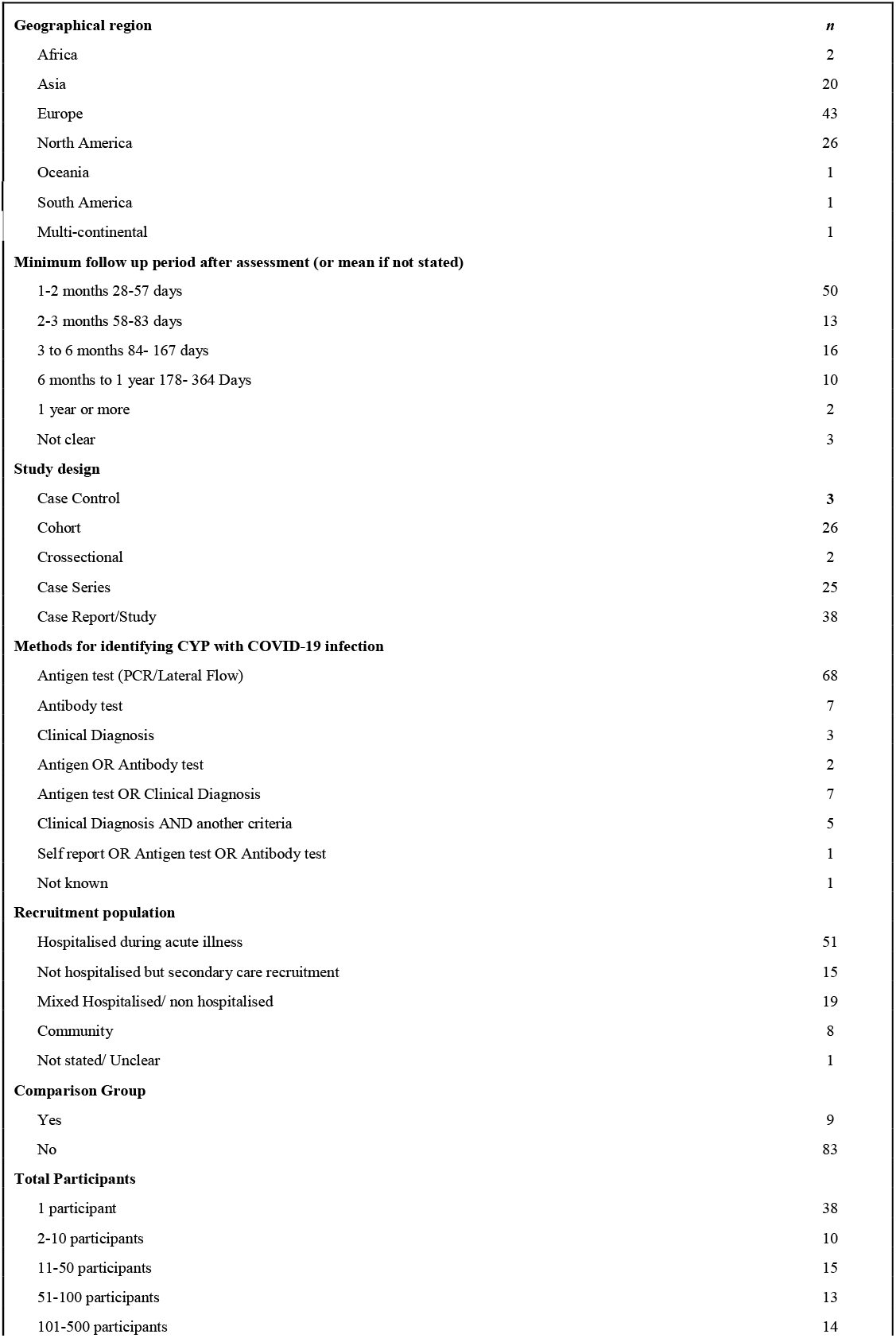
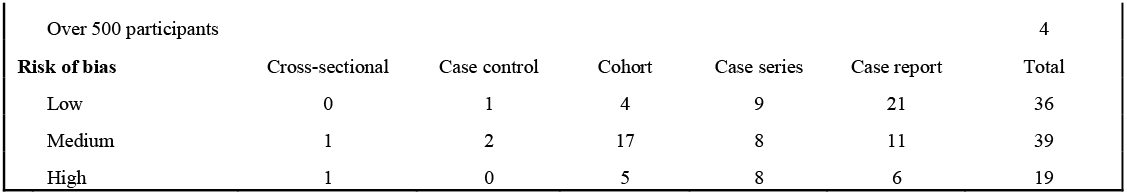
Summary of study characteristics.

|  |  |
| --- | --- |
| **Geographical region** | ***n*** |
| Africa | 2 |
| Asia | 20 |
| Europe | 43 |
| North America | 26 |
| Oceania | 1 |
| South America | 1 |
| Multi-continental | 1 |
| **Minimum follow up period after assessment (or mean if not stated)** |  |
| 1-2 months 28-57 days | 50 |
| 2-3 months 58-83 days | 13 |
| 3 to 6 months 84- 167 days | 16 |
| 6 months to 1 year 178- 364 Days | 10 |
| 1 year or more | 2 |
| Not clear | 3 |
| **Study design** |  |
| Case Control | 3 |
| Cohort | 26 |
| Crossectional | 2 |
| Case Series | 25 |
| Case Report/Study | 38 |
| **Methods for identifying CYP with COVID-19 infection** |  |
| Antigen test (PCR/Lateral Flow) | 68 |
| Antibody test | 7 |
| Clinical Diagnosis | 3 |
| Antigen OR Antibody test | 2 |
| Antigen test OR Clinical Diagnosis | 7 |
| Clinical Diagnosis AND another criteria | 5 |
| Self report OR Antigen test OR Antibody test | 1 |
| Not known | 1 |
| **Recruitment population** |  |
| Hospitalised during acute illness | 51 |
| Not hospitalised but secondary care recruitment | 15 |
| Mixed Hospitalised/ non hospitalised | 19 |
| Community | 8 |
| Not stated/ Unclear | 1 |
| **Comparison Group** |  |
| Yes | 9 |
| No | 83 |
| **Total Participants** |  |
| 1 participant | 38 |
| 2-10 participants | 10 |
| 11-50 participants | 15 |
| 51-100 participants | 13 |
| 101-500 participants | 14 |

|  |  |  |  |  |  |  |
| --- | --- | --- | --- | --- | --- | --- |
| Over 500 participants |  |  |  |  |  | 4 |
| **Risk of bias** | Cross-sectional | Case control | Cohort | Case series | Case report | Total |
| Low | 0 | 1 | 4 | 9 | 21 | 36 |
| Medium | 1 | 2 | 17 | 8 | 11 | 39 |
| High | 1 | 0 | 5 | 8 | 6 | 19 |

Included studies came from 47 different countries although the majority were from Europe (43)^24,28-32,34,36,39,40,44,49,50,52,53,55,58-60,68,74,75,77-79,81,82,84,88-91,94-96,98,103,104,107,113,114,^and the USA (26)^33,35,37,41-43,45-48,57,61,63,67,69-71,76,83,86,93,99,109-112^. Articles were translated from Norwegian, Russian, and Mandarin. 38 were case reports^23,33,36,41,43,47,48,51,52,55,57,60-62,64,66-70,72,78,80,85-87,89,92,98-101,107,108,110,112^. Of those that were not case reports, most were cohort studies (26)^23,24,33,36,41,43,47,48,51,52,55,57,60-62,64-70,72,78,80,85-87,89,92,98-101,107,108,110,112^. Most studies recruited from hospital or secondary care settings, eight recruited solely from community settings^38,81,82,90,91,94,107,114^.

Most studies only included participants who had had a positive antigen test for Sars-CoV-2, although 16 studies included participants where diagnosis was either clinical or by self-report^31,35,42,70,73,74,75,80,81,83,98,102,106,113,115,116^. The longest duration of follow up was 10-13 months^79^. 36 studies were judged to have low risk of bias^23,27,28,32,33,36,38,41,46,48,51,52,55,58,59,64-66,69,71,73,76,78,81,83,85-87,89,94,98,100,109,110,112,114,^ with 19 studies in the high-risk group.^25,30,35,39,47,53,54,57,62,75,77,79,80,92,99,102,105,111,115^

Of 26 cohort studies^26,29,30,37,39,40,44,50,53,54,56,63,76,81,82,88,90,91,94-97,103,104,114^, 17 were judged as moderate risk of bias^26,29,37,40,44,50,56,63,82,88,90,91,95-97,103,104^. The commonest bias risks were the identification and handling of potential confounding factors, and lack of generalisability, often due to small sample size. The four cohort studies deemed low risk were large, matched cohorts^37,81,94,114^. Of 25 case series^25,27,28,31,32,34,35,42,46,59,71,73-75,77,79,83,93,102,106,109,111,113,115,116^, a third were high^25,35,75,77,79,102,111,116^, a third moderate^31,34,42,74,93,106,113,115^ and a third low risk of bias^27,28,32,46,59,71,73,83,109^; Lack of detail on completeness of inclusion and the clinical setting / demographics of participants were the commonest bias risks.

### 1) Patterns of symptoms lasting longer than four weeks

Where symptoms of COVID-19 continued for more than four-weeks, more than 100 symptoms affecting multiple body systems were reported (Figure 2). Fatigue was reported in the highest number of studies (478 participants out of 1192 with prolonged symptoms in 23 studies), followed by headache (192/1121/18) and cognitive symptoms (248/949/15). After constitutional and neurological symptoms, the most affected systems were respiratory, ear, nose and throat, and musculoskeletal.

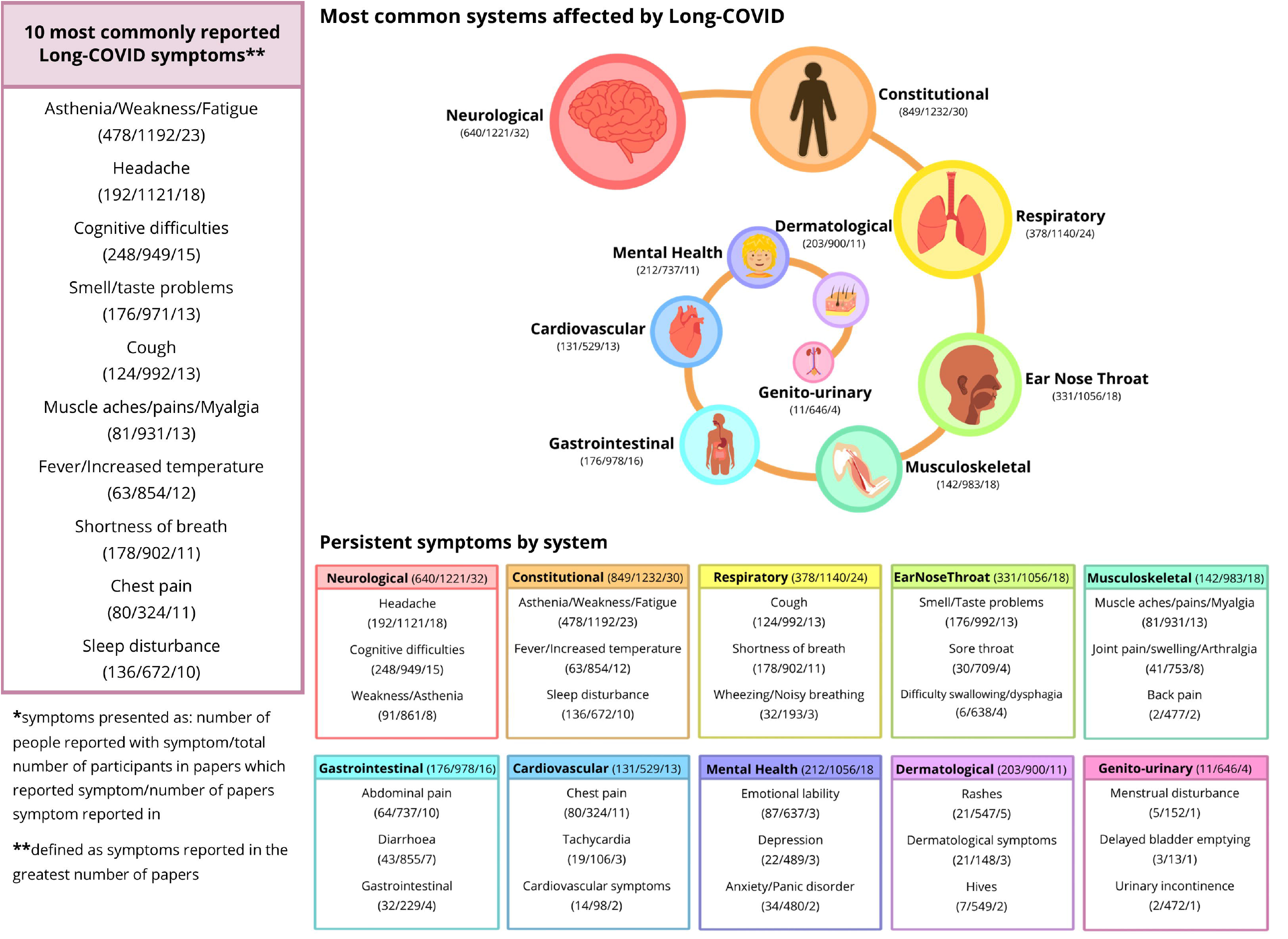

### 2) Risk of developing prolonged symptoms

17 studies^26,30,32,53,56,74,79,81,82,88,91,94,95,97,103,104,114^ (Table 2) reported numbers of participants with prolonged symptoms from a defined population of participants with COVID-19 and were included in a meta-analysis. 11 of these recruited from hospital settings^26,30,32,53,56,79,88,95,97,103,104^ and six recruited from the community^74,81,82,91,94,114^. 14 were cohort studies^26,30,53,56,81,82,88,91,94,95,97,103,104,114^, and three were longitudinal case series^32,74,78^ (all three case series contained at least 70 participants).

**Table 2:** Studies included in the quantitative synthesis.

| Author | Setting | Duration of prolonged symptoms | Cases |  |  | Controls |  |  |
| --- | --- | --- | --- | --- | --- | --- | --- | --- |
|  |  |  | Definition of n | Definition of N | n/N | Definition of n | Definition of N | n/N |
| Molteni et al. (2021) | Community | Beyond 4 weeks | Seropositive and calculable illness duration and at least 1 persisting symptom | Matched control group 1:1 | 77/1734 | Seronegative and calculable illness duration and at least 1 persisting symptom | Matched control group 1:1 | 15/1734 |
| Miller et al. (2022) | Community | Beyond 4 weeks | At least 1 persisting symptom and evidence of past or present covid infection | All participants with evidence of past or present COVID infection | 43/1062 | Persisting symptoms but no evidence of past or present COVID infection | All participants with no evidence of past or present COVID infection | 86/3970 |
| Radtko et al. (2021) | Community | Beyond 4 weeks | Seropositive with prolonged symptoms | All seropositive participants | 10/109 | Seronegative with prolonged symptoms | All seronegative participants | 121/1246 |
|  |  | Beyond 12 weeks |  |  | 4/109 |  |  | 28/1246 |
| Roge et al. (2021) | Hospital | Beyond 12 weeks | Seropositive with at least 1 persisting symptom | All seropositive participants | 152/236 | Other viral illness with at least one persisting symptom | All participants in with other viral illness | 32/142 |
| Zavala et al. (2021) | Community | Beyond 4 weeks | Seropositive with prolonged symptoms | All seropositive participants | 24/472 | Seronegative with prolonged symptoms | Seronegative without prolonged symptoms | 6/387 |
| Asadi-Pooya et al. (2021) | Hospital | Beyond 12 weeks | Seropositive with prolonged symptoms | Total participants | 26/58 |  |  |  |
| Bossley et al. (2021) | Hospital | Beyond 4 weeks | At least 1 persisting symptom and previous admission with covid | Total children admitted and followed up | 11/71 |  |  |  |
| Buonsenso et al. (2021) | Hospital | Beyond 4 weeks | Seropositive with prolonged symptoms | Total participants | 75/129 |  |  |  |
| Erol et al. (2021) | Hospital | Beyond 4 weeks | Seropositive with prolonged symptoms | Total participants | 45/121 |  |  |  |
| Fink et al. (2021) | Hospital | Beyond 4 weeks | Seropositive with prolonged symptoms | Total participants | 23/53 |  |  |  |
|  |  | Beyond 12 weeks |  |  | 12/53 |  |  |  |
| Osmanov et al. (2021) | Hospital | Beyond 12 weeks | Seropositive with prolonged symptoms | Total participants | 128/518 |  |  |  |
| Matteudi et al. (2021) | Hospital | Beyond 12 weeks | Seropositive with prolonged symptoms | Total participants | 23/137 |  |  |  |
| López et al. (2021) | Community | Beyond 4 weeks | Presumed COVID illness with persistent symptoms at follow up | Total participants | 8/72 |  |  |  |
| Powell et al. (2021) | Community | Beyond 4 weeks | Seropositive with prolonged symptoms | All seropositive participants | 7/259 |  |  |  |
| Say et al. (2021) | Hospital | Beyond 4 weeks | Seropositive with prolonged symptoms | Total participants | 12/151 |  |  |  |
| Smame et al. (2020) | Hospital | Beyond 12 weeks | Seropositive with prolonged symptoms | Total participants | 9/30 |  |  |  |
| Sterky et al. (2021) | Hospital | Beyond 12 weeks | Seropositive with prolonged symptoms | Total participants | 12/55 |  |  |  |

\*hospital includes studies of admitted patients and mixed in-patient / outpatient recruitment

Summary estimates of risk of prolonged symptoms were higher hospital samples (33·9%, 95% CI 21·5% to 47·4%) than in community samples (5·1%, 95% CI 3·6% to 7·0%) (Figure 3). Studies were stratified by those reporting symptoms beyond 12 weeks and those reporting symptoms between 4 and 12 weeks. Studies with longer follow-up reported higher rates of prolonged symptoms (27%, 95% CI 13·3% to 43·2%) than those with short follow-up (14·6%, 95% CI 8·1% to 22·6%) (Figure 4). In these subgroup analyses, the estimate of between study heterogeneity (**τ**2) varied from 0·111 (community population) to 0·224 (duration beyond 12 weeks).

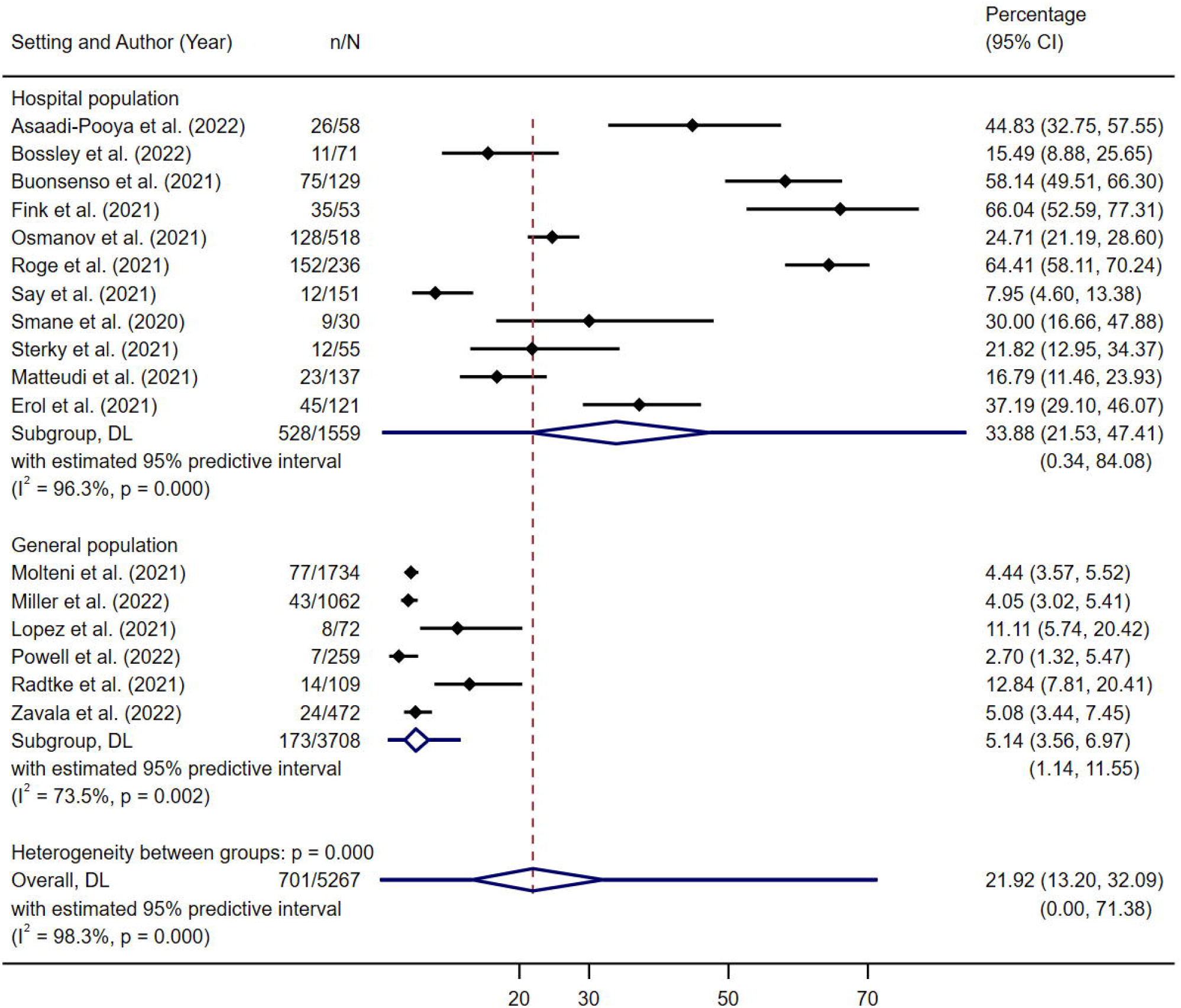

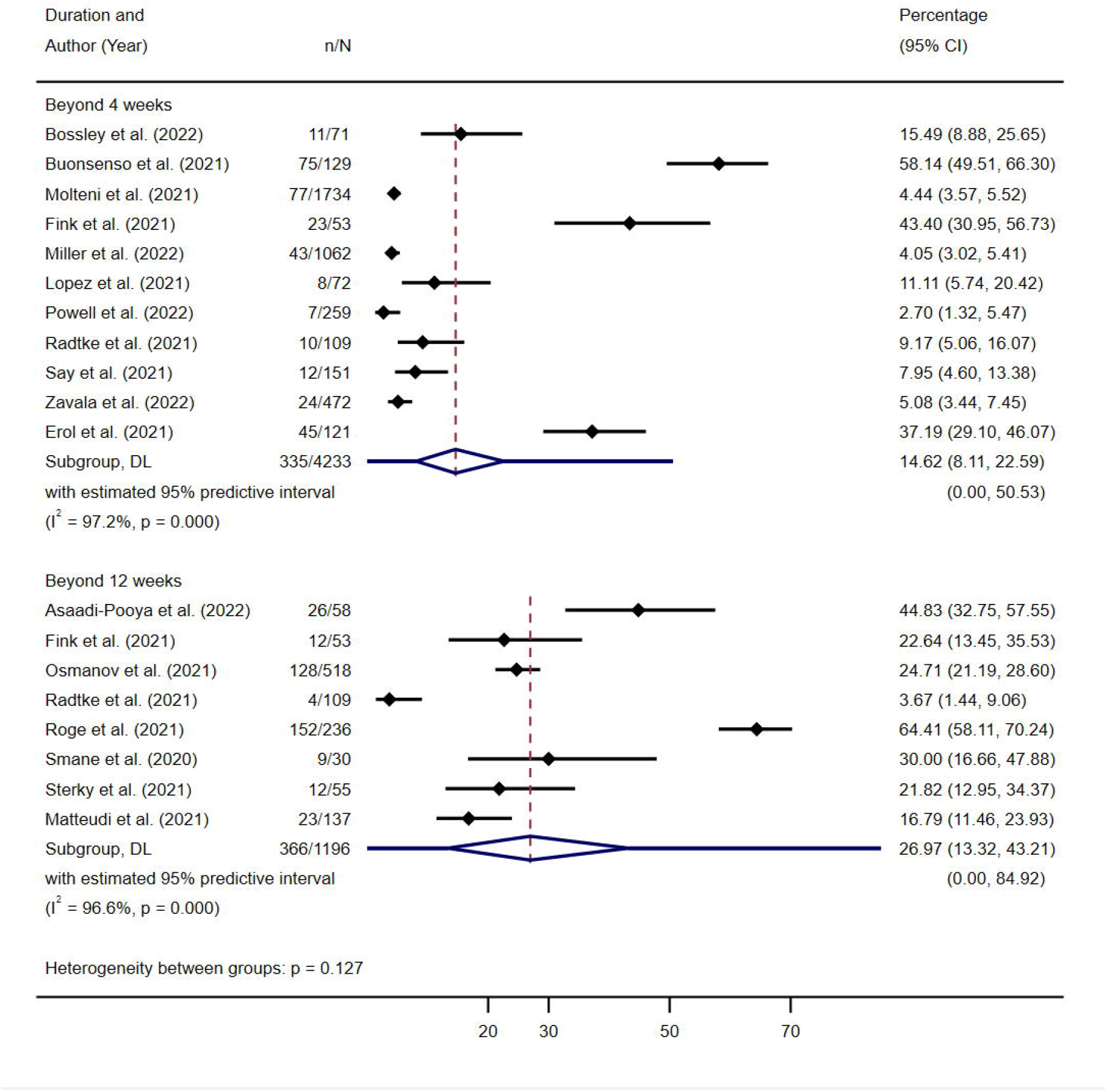

Five studies reported comparator data^81,82,94,95,114^. Four were set in the community and controls were sero-negative or had no evidence of past infection^81,82,94,114^. One recruited from a post-acute hospital clinic (participants included those hospitalised and not-hospitalised for the acute illness) and the control group comprised children with other viral illnesses^95^. In the community studies, the estimated summary OR of prolonged symptoms in the COVID-19 positive group compared to the non-COVID group was 2.4 (95% CI 1·2 to 4·7) (Figure 5), indicating higher likelihood of prolonged symptoms in CYP with COVID-19 compared those without COVID-19. This result is subject to substantial between-study heterogeneity (*τ*2 0·372, *I*2 82%, *P*<0·001). We assessed publication bias for risk of prolonged symptoms in the studies recruiting from hospital (the community group contained <10 studies). Funnel plot asymmetry showed potential small-study effects on funnel plot (Supplementary data, S4) although this was not statistically significant (β = 5·4, t = 1·36, P = 0·206).

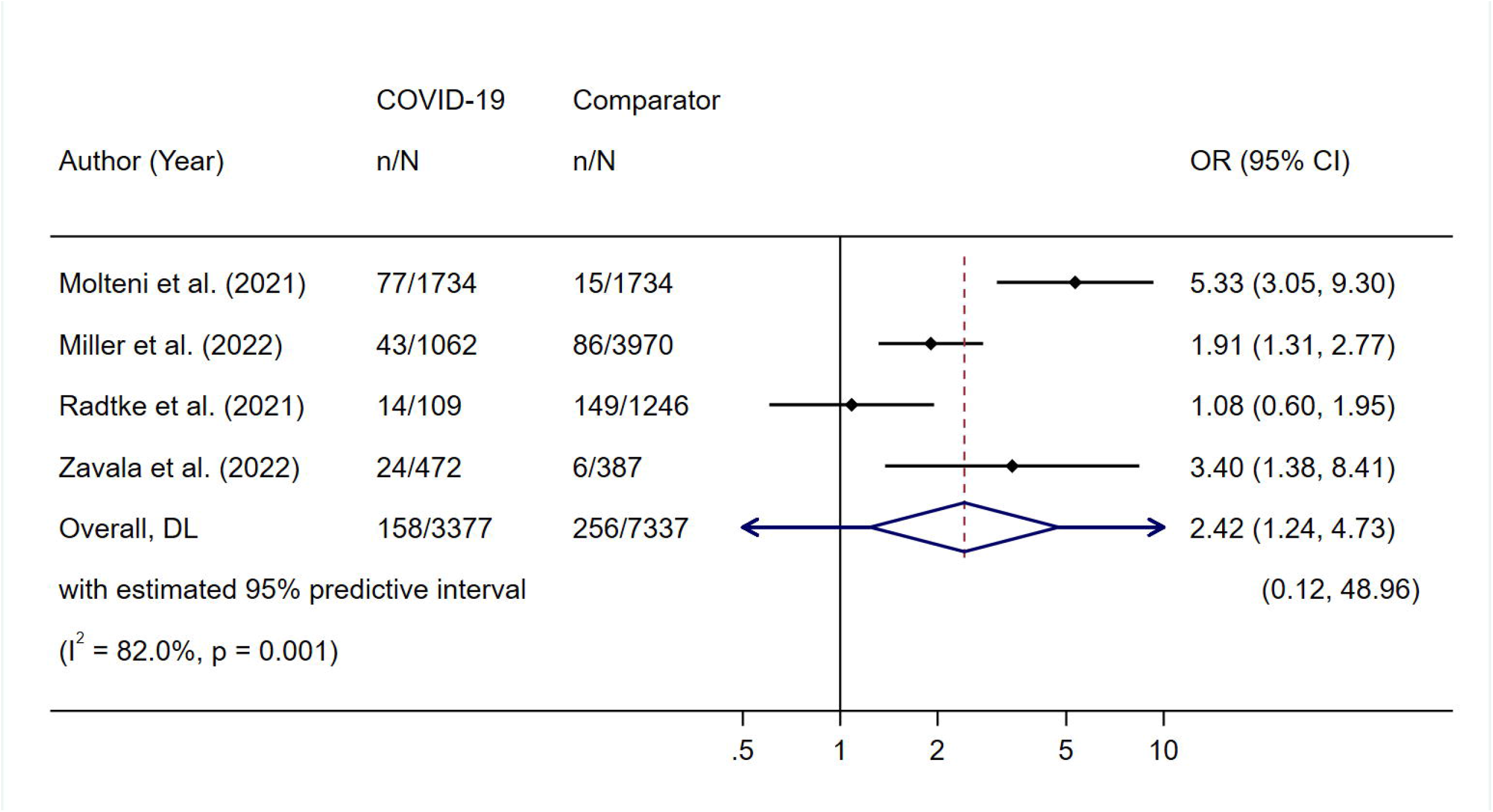

Two cohort studies^39,90^ reported group level findings rather than individual numbers of participants affected and thus were not included in the meta-analysis. Clavenna,^39^a matched cohort study deemed high risk of bias, reported that the prevalence of symptoms over six months of follow up was similar in those who had had COVID-19 and those who had not. Petersen^90^, deemed moderate risk of bias, reported that children were less likely than adults to experience prolonged symptoms after COVID-19.

### 3) Sequelae of SARS-CoV-2 infection (including clinical effects and persistent radiological and pathological findings)

17 studies (two cohort studies^29,37^, four case series^71,102,106,109^, one cross-sectional study^38^ and 10 case studies^24,47,61,64,65,80,86,87,98,100^) described clinical sequalae of COVID-19 occurring after, or lasting more than, four-weeks from the acute infection. One large, matched cohort study (predominantly adults but included 2673 matched pairs of children), deemed at moderate risk of bias^37^, observed all new conditions presenting one to four months after COVID-19 and reported that children who had had COVID-19 were not more likely to develop new conditions than controls.

Stroke was reported as an outcome in four studies: one case series^71^, three single case studies^65,98,100^. The case series, assessed as low risk of bias, observed three cases of acute ischaemic strokes, and four cases of haemorrhagic stroke amongst 1695 children hospitalised due to COVID-19.

The association between new-onset diabetes and COVID-19 was considered in five studies. Two looked at populations of children presenting to hospital with type-1 diabetes (Trieu^109^, n=576; Boboc^29^, n=147) for evidence of co-existent SARS-CoV-2 infection. Both studies were considered as low risk of bias and found nine cases of COVID-19 in 433 new cases of type-1 diabetes^,109^, and one case of COVID-19 in 290 new onset type-2 diabetes combined^109^. One case series (Thakur & Rai^106^, n=2), and two case studies^86,87^ also described new cases of new onset type-1 diabetes with recent COVID-19.

Steroid-dependant autoimmune haemolytic anaemia was reported in one case from 397 children with confirmed SARS-CoV-2 infection^38^. Other sequelae reported in case series or case studies include acute pancreatitis with pseudocyst formation^102^, Guillain-Barre syndrome (3 case studies^24,64,80^, acute psychosis^61^ (one 17-year-old), and concurrent rheumatic fever^47^ (one case study).

Ten studies reported laboratory or radiological abnormalities that persisted for more than four-weeks from COVID-19 infection despite symptom resolution^44,57,58,66,68,78,92,99,105,115^. Two cohort studies reported persistent abnormal lung imaging in children previously hospitalised with COVID-19 (22 cases in Tang^105^, n=46, high risk of bias; and ten in Denina^44^, n=25, moderate risk of bias). A case series (n=14) also reported persisting chest CT abnormalities in seven children^115^. Retinal vasculature abnormalities were found in children who had COVID-19 in a case-control study (cases=63) deemed low risk of bias^58^. Six case studies reported other abnormal blood or imaging findings persisting beyond four-weeks, including leucopaenia^68,92,99^, idiopathic thrombocytopaenia^68^, raised inflammatory markers^44^, right coronary artery dilatation on echocardiogram^57^, and residual spinal MRI changes after clinically resolved Guillan-Barre syndrome associated with COVID-19^66^.

### 4) Effects in children with pre-existing conditions

16 studies (including nine case studies) described the effects of COVID-19 in children with specified underlying conditions and reported prolonged symptoms or complications in at least some of the participants^23,27,28,33,42,48,51,59,60,62,67,72,76,89,110,113^. The underlying conditions were predominantly associated with immunodeficiency, either due to the primary condition or its treatment.

Only three of these studies were cohorts or large case series. Madhusoodhan^76^ was a multi-centre study including 98 children with malignancies who tested positive for Sars-CoV-2. Reported range of symptoms was up to 52 days (median 10) and range of duration of in-patient care for the acute illness was up to 65 days (median 12). Kamdar^63^ followed 109 children with haematological-oncological conditions who had COVID-19 and reported only one child with a late complication, although duration of symptoms for most participants was not reported. Conway^42^ was a case series of 225 heart transplant candidates or recipients who had COVID-19, in which seven had on-going symptoms at 30 days.

Other studies in this category were small case series or case reports. Most described prolonged COVID-19 illness complicated by the underlying condition or its treatment^27,33,48,67,72,89,110,113^. Some described complications that were thought to be due to COVID-19 such as myocarditis occurring two months after COVID-19 in a child with renal failure^60^, right atrial thrombus in a child with acute lymphoblastic leukaemia (ALL)^23^ and immune thrombocytopaenic purpura occurring 22 days after COVID-19 in a child with ALL^51^. Others described an effect of COVID-19 on the underlying condition e.g. three children with flares of juvenile idiopathic arthritis occurring or lasting more than four weeks after COVID-19^59^.

## Discussion

This review describes the spectrum of reported outcomes beyond four-weeks in children who have had COVID-19. The most reported prolonged symptoms were fatigue, headache and cognitive difficulties. Summary estimates of risk of prolonged symptoms were higher in studies recruiting from hospital settings (31·2%, 95% CI 20·3% to 43·2%) than community settings (4·6%, 95% CI 3·4% to 5·8%). Children who had had COVID-19 were more likely (OR 2·96) to experience prolonged symptoms than those who had not. Sequalae including stroke, type-1 diabetes, Guillan-Barre syndrome, and persistent radiological or blood test abnormalities have been reported in CYP following COVID-19 but most studies reporting these are case reports / case series and quality of evidence is low.

Like previous reviews, we found that where COVID-19 symptoms persist for more than four-weeks in children, they are multisystemic and vary widely between individuals. In alignment with our findings, two previous systematic reviews of persistent symptoms of COVID-19 in CYP^11,12^ also found constitutional and neurological symptoms to be amongst the most reported. This is analogous to studies of adults with Long-COVID where systematic reviews^117,118^ have found general malaise, fatigue, sleep disturbance and concentration impairment to be commonly reported. In adult populations however, breathlessness and altered sense of smell are often higher up the ranking of symptom frequency^14,117,118,119^.

Most studies in the meta-analysis of prolonged symptom prevalence included children recruited from hospital settings. However, stratification by setting showed that risk of prolonged symptoms was higher in studies of children recruited from hospital settings than in studies recruiting from community settings. This is important, given most children with COVID-19 have mild illness and do not require hospitalisation. There have been large community studies published since the search window for this review, which may go some way to redressing this balance. These include the CLoCK study, a matched cohort study of post-COVID-19 condition among children and adolescents 11 to 17 years of age that recruited participants using the British national testing database^10^ and a further matched cohort study from Germany^14^ which used routine health care data to report outcomes at least three months after COVID-19.

Rates of prolonged symptoms in the studies included in the meta-analysis was higher in studies that reported outcomes beyond 12 weeks than in those that reported symptoms at four-weeks. This is to be interpreted with caution as the numbers in the studies with longer follow-up were smaller and there was only one study in this group which recruited from a community setting. Whilst the NICE and United States Centres for Disease Control and Prevention (CDC) definitions of post-COVID conditions encompass symptoms continuing or occurring more than four-weeks after acute infection, the World Health Organisation (WHO) has recently published a consensus definition of Post COVID-19 condition in children and adolescents, which refers to “individuals with a history of confirmed or probable SARS-CoV-2 infection, experiencing symptoms lasting at least 2 months which initially occurred within 3 months of acute COVID-19”^120^. This review highlights that the evidence base for characterising and understanding symptoms over this duration in CYP is sparse.

Only five studies included a comparator group. Given the broader effects of the pandemic and associated social restrictions on children’s health, high quality matched studies are critically important to understanding effects directly attributable to Sars-CoV-2 infection. From this review’s limited data, the likelihood of having symptoms lasting more than four-weeks is higher in children who have had COVID-19 than in those that have not. Of note, both large community studies mentioned above included a control group. The CLoCK study’s^10^ first follow-up data showed that children with a positive test were significantly more likely to report multiple symptoms at three months than controls, and Roessler^14^ found increased rates of physical and mental health conditions in cases than controls, with highest incident rate ratios for malaise, fatigue/exhaustion and cough.

Children with medical complexity and certain underlying conditions have been shown to be at higher risk of severe COVID-19^121^. The studies in this review that focussed on children with specific conditions did not demonstrate high numbers of children experiencing long-term effects. However, there were only three cohorts / case series in this group and they were not designed to study long-term outcomes, hence firm conclusions cannot be drawn.

Our review is unique in that, alongside pattern and risk of prolonged symptoms, we have synthesised evidence on other sequalae of COVID-19 in CYP. The most common conditions reported to have developed in temporal association with COVID-19 were neurological conditions, including stroke and Guillain-Barre syndrome, and type-1 diabetes. In adults, COVID-19 infection has been associated with a range of long-term conditions / complications including new onset diabetes^122,123^ venous thromboembolism^14^, neurological complications including stroke^124,125^ and cardiovascular disease^126^. The extent to which CYP are affected by similar sequalae is less well characterised. One recent report using US CDC data^121^ did identify increased risk of acute pulmonary embolism (adjusted hazard ratio = 2·01), myocarditis and cardiomyopathy (1·99), venous thromboembolic event (1·87), acute and unspecified renal failure (1·32), and type-1 diabetes (1·23) in under 18s, but this needs further confirmation.

Synthesis of persisting radiological and pathological findings following COVID-19 in CYP is also novel. A systematic review of long-term effects of COVID-19 in adults looked at persistently abnormal clinical investigations in hospitalised patients and found lasting changes in lung function and structure^118^ but previous reviews in paediatric populations have not considered this. The frequency, clinical significance and duration of these persisting abnormalities remains unknown and warrants further exploration.

The main limitation of this study was the time taken to complete the work set against the rapid increase in the number of studies being published. Most studies were from high income countries. Heterogeneity in diagnosis of COVID-19, classification of symptoms and the method and duration of follow up all made synthesis less secure. Risk of attribution bias in symptom reporting and low numbers of studies with control groups also limits the ability to draw firm conclusions. Assessment of publication bias was only possible in studies recruiting from hospital settings, and analysis showed potential small study effects in this group. Few studies reported results stratified by age and there was no data on differences in patterns or prevalence of symptoms in different ethnicities, both of which need consideration in developing age and culturally appropriate resources and services.

This review adds to the evidence that some CYP experience effects of COVID-19 that last longer than four-weeks, and describes the most common prolonged symptoms, risk of prolonged symptoms and broader sequalae of the acute infection. It also highlights gaps in the evidence. To develop treatment plans for affected CYP and to plan appropriate services to support them, further studies are needed to better characterise this condition. These should ideally recruit from community settings, include population-based control groups and use standardised definitions and outcome measures where possible.

## Data Availability

Full extracted data and risk of bias assessment results will be available on request.

## Contributors

CB, VW, HT and NC developed the protocol and NC carried out the searches.

CB, VW, HT, GS, TS and AF did the screening and data extraction and have accessed and verified the data.

RB did the statistical analysis.

All authors contributed to the writing of the manuscript.

## Declarations of interest

We declare no competing interests.

## Data sharing

Full extracted data and risk of bias assessment results will be available on request.

## Acknowledgements

There was no grant funding specifically for this review, but authors are supported by the funding streams previously stated. We wish to acknowledge our translators, Anthony Burton and Dahai Yu.

## Notes

### Competing Interest Statement

The authors have declared no competing interest.

### Funding Statement

There was no grant funding specifically for this review, but HT, CB and GS have National Institute for Health and Care Research fellowships. RB, CM and VW are supported by the NIHR West Midlands Applied Research Collaboration. CM Is supported by the NIHR School for Primary Care Research.

