## Supplementary material for "Long-term outcomes of COVID-19 infection in children and young people: a systematic review and meta-analysis"

Supplementary material – S1

| Author | Report type | Country | Study aim | Setting / severity of acute COVID-19 | Age (years) | Gender %F | COVID-19 diagnosis* | Duration of follow up / timepoint of assessment following acute illness | Cases | | Comparator group | | Risk of bias** |
| --- | --- | --- | --- | --- | --- | --- | --- | --- | --- | --- | --- | --- | --- |
|  |  |  |  |  |  |  |  |  | Total | n with prolonged symptoms / effects | Total | n with prolonged symptoms / effects |  |
| Prolonged symptoms | | | | | | | | | | | | | |
| Ludvigsson (2020) | Case series | Sweden | Described five cases of children with long COVID-19, based on parental reports. | Non-hospitalised | Median 12 (range 9-15) | 80% | Cl | 8 months | 5 | 5 |  |  | H |
| Lopez et al. (2021) | Case series | Spain | Described persisting symptoms identified during the routine follow up of children with an initial COVID-19 diagnosis | Non-hospitalised | Median 12 (IQR 10-14) | 50% | Cl or Ag | Median 53 (25–61) days | 72 | 8 |  |  | M |
| Buonsenso et al. (2021) | Case series | Italy | Described persisting symptoms in paediatric patients previously diagnosed with COVID-19 | Mixed hospitalised / non-hospitalised | Mean 11 (SD 4.4) | 48% | Ag | Mean 162 days | 129 | 75 |  |  | L |
| Smane et al. (2020) | Retrospective cohort | Latvia | Identified data describing persisting symptoms after recovery from COVID-19 in children | Mixed hospitalised / non-hospitalised | Mean 9 (SD 5.2) | 43% | Ag | Mean 101 days | 30 | 9 |  |  | M |
| Osmanov et al. (2021) | Prospective cohort | Russia | Assessed the longterm outcomes of children previously hospitalised with COVID-19, and associated risk factors | Hospitalised | Median 10 (range 0-18) | 52% | Ag | Range 5-9 months | 518 | 128 |  |  | M |
| Di Sante et al. (2021) | Case control | Italy | Described immunological differences between children with post-acute sequelae of SARS-CoV-2 and those who made a full recovery | Mixed hospitalised / non-hospitalised | Mean 10 (SD 4.5) | 35% | Ag | 5+ weeks | 29 | 12 |  |  | M |
| Zhvania  et al. (2021) | Case series | Georgia | Presented clinical observations of young patients following an episode of COVID-19 | Mixed hospitalised / non-hospitalised | Range 0-17 | 45% | Cl plus Ab | 2+ months | 60 | 60 |  |  | H |
| Brackel et al. (2021) | Case series | Netherlands | Determined the size of the pediatric population experiencing symptoms of long-COVID, who had been referred to a specialist and illustrated and identified specific disease characteristics | Mixed hospitalised / non-hospitalised | Median 13 (IQR 9-15, range 2-18) | Unknown | Cl or Ag | Months' | 89 | 89 |  |  | M |
| Powell et al. (2021) | Cohort | UK | Described the national epidemiology, risk factors, clinical features, and outcomes of SARS-CoV-2 infection in primary school aged children after the partial reopening of schools in England | Community testing | Median 7 (IQR 5-10) | 47% | Ag | 1 month | 259 | 7 |  |  | M |
| Sterky et al. (2021) | Cohort | Sweden | Assessed the extent, and type of persistent symptoms in children admitted to two paediatric hospitals due to Covid-19 | Hospitalised | 0-18 years | Unknown | Ag | 4+ months | 55 | 12 |  |  | M |
| Say et al. (2021) | Cohort | Australia | Described the medium-term clinical outcomes 3-6 months after diagnosis in children with COVID-19 presenting to a tertiary paediatric hospital | Mixed hospitalised / non-hospitalised | Median 3 (IQR 1-8] | 47% | Ag | 3-6 months | 171 | 12 |  |  | M |
| Fink et al. (2021) | Cohort | Brazil | Prospectively assessed the demographic, anthropometric, clinical and health-related quality of life data in paediatric patients with COVID-19 | Mixed hospitalised / non-hospitalised | Median 15 (range 8-18) | 59% | Ag | Median 4.4 months | 53 | 23 |  |  | M |
| Morrow et al. (2021) | Case series | USA | Described a case series of 9 paediatric patients attending a post COVID-19 rehabilitation clinic | Mixed hospitalised / non-hospitalised | Median 13 (range 4-18) | 67% | Cl or Ag | 2+ months | 9 | 9 (1 MIS-C) |  |  | L |
| Esmaeilzadeh et al. (2022) | Cohort | Iran | Described the association of asthma-like symptoms in hospitalised children affected by COVID-19 and examined whether hospitalisation because of COVID-19 resulted in the development of persistent cough and asthma-like symptoms | Hospitalised | 0-18yrs old | 39% | Ag | 6 months | 69 | 27 |  |  | H |
| Bossley et al. (2021) | Cohort | UK | Explored whether children with SARS-CoV-2 RNA positivity might have late symptoms in common with post-acute covid-19 syndrome in adults | Hospitalised | Range 0-17 | 39% | Ag | 3+ months | 88 | 11 |  |  | H |
| Clemente et al. (2021) | Cohort | Italy | Described experience of telemedicine and presented results of the follow up of 65 children admitted due to Covid | Hospitalised | Range 0-16 | 32% | Ag | 1+ months | 19 | 6 |  |  | M |
| Miller et al. (2022) | Cohort | UK | Estimated the prevalence of persistent symptoms reported among children, including those with a history of COVID-19 infection. Identified risk factors associated with persistent symptoms during the COVID pandemic among all children participating in a large household-based community cohort study. | Community recruitment - treatment setting unclear | Range 0-17 | Unknown | Self report or Ag or Ab | 1+ months | 1062 | 43 | 3970 | 86 | L |
| Castelo-Soccio et al. (2021) | Case series | USA | Described a cohort of children with acral changes with the aim of understanding an association with SARS-CoV-2 | Non-hospitalised | Mean 13 (range 0-18) | 39% | Cl | 1+ months | 114 | 114 |  |  | H |
| Lindan et al. (2020) | Case series | Multi- national | Evaluated neuroimaging manifestations of COVID-19 in the paediatric population | Hospitalised | Range 0-16 | 45% | Cl or Ag | 1+ months | 38 | 11 |  |  | L |
| Dolezalova et al. (2021) | Cohort | Czechia | Explored the clinical picture, severity, and prognosis of post-COVID syndrome in children with a focus on the respiratory system | Hospitalised | Median 14 (IQR 8-15) | 56% | Ag | 6 months | 39 | 34 |  |  | M |
| Calitri et al. (2021) | Case series | Italy | Described the COVID-19 course in a population of children from diagnosis to possible hospital admission and recovery, with a long-term clinical and serological follow-up | Mixed hospitalised / non-hospitalised | Median 8 (range 0-14) | 52% | Ag | 1+ months | 46 | 1 |  |  | M |
| Alshengeti et al. (2021) | Case series | Saudi Arabia | Described COVID-19 among children in Al-Madinah, Saudi Arabia | Hospitalised | Mean 3 (range 0-13) | 51% | Ag | 1+ months | 106 | 1 |  |  | H |
| Venn et al. (2020) | Case series | USA | Described three cases of croup in children with COVID-19 infection who received nebulised racemic epenephrine in an emergency department setting | Hospitalised | Median 2 (range 0-9) | 66% | Ag | 2 months | 3 | 1 |  |  | H |
| Zavala et al. (2021) | Matched cohort | UK | Determined the course of illness and ongoing symptoms in children with laboratory-confirmed SARS-CoV-2 infection compared with test-negative children in England during Jan 2021 when Alpha variant was prevalent | Community testing | Median 10 (IQR 6-13) | 49% | Ag | 1+ months | 472 | 24 | 387 | 6 | L |
| Molteni et al. (2021) | Matched cohort | UK | Described illness duration, individual symptom prevalence and duration, and symptom burden in UK school-age children testing positive for SARS-CoV-2, and provide similar data for symptomatic children testing negative during the same period. | Community recruitment | Median 13 (IQR 10-15) | 50% | Ag | 1+ months | 1734 | 77 | 1734 | 15 | M |
| Clavenna et al. (2021) | Cohort | Italy | Described the characteristics of children visited for a suspected SARS-CoV-2 infection and to monitor their health status in the 6 months after the first visit. | Mixed hospitalised / non-hospitalised | Median 7 (IQR 4-11) | 50% | Ag | 6 months | 41 | Unclear | 107 | Unclear | H |
| Roge et al. (2021) | Cohort | Latvia | Identified the long-term consequences of SARS-CoV-2 infection in children and compared the persistent symptom spectrum between COVID-19 and community-acquired infections of other etiologies. | Mixed hospitalised / non-hospitalised | 0-18 years | 45% | Ag | 1-6 months | 236 | 152 | 142 | 32 | M |
| Radtke et al. (2021) | Cohort | Switzerland | Compared long COVID compatible symptoms in children with 6-months follow-up according to their SARS-CoV-2 serology | Community recruitment | Range 6-16 | 46% | Ag | 6 months | 109 | 10 | 1246 | 121 | L |
| Petersen et al. (2021) | Cohort | Faroe Islands | Described symptoms during the acute phase and long Covid symptoms in patients from the Faroe Islands | Community recruitment | Range 0-17 | Unknown | Ag | Mean 125 days (SD 18) | 21 | Unclear |  |  | M |
| Denny et al. (2021) | Cross-sectional | USA | Describe the severity and clinical course of COVID-19 disease in children, identified clinical risk factors for severe disease. | Non-hospitalised | Median 16 (IQR 9-19) | 53% | Ag | 1+ months | 40 | Unclear |  |  | M |
| Reiff et al. (2021) | Case series | USA | Presented the experiences of COVID-19 and MIS-C at the authors' institution and compared the two disease processes Jersey. | Hospitalised | Mean 16 (IQR 9-17) | Unknown | Ag or Ab | Duration of admission (up to 47 days) | 90 | Unclear |  |  | M |
| Derespina et al. (2020) | Case series | USA | Described the manifestations of critically ill children with COVID-19 admitted to pediatric intensive care units (PICUs) across New York City during the first wave of the US pandemic and to identify factors associated with PICU and hospital length of stay (LOS). | Hospitalised | Median 15 (IQR 9-19) | 39% | Ag | 28 days | 70 | 14 |  |  | L |
| Matteudi et al. (2021) | Case series | France | Described acute symptoms and long term consequences of children testing postitive for COVID-19 in Marseille | Mixed hospitalised / non-hospitalised | Mean 9 (Range 0-16) | Unknown | Ag | 10-13 months | 137 | 23 |  |  | H |
| Rusetsky et al. (2021) | Cohort | Russia | Evaluated the olfactory status in children with laboratory confirmed SARS‐CoV‐2 using subjective and psychophysical methods | Mixed hospitalised / non-hospitalised | Median 13 (range 5-17) | 53% | Ag | 2 months | 79 | 3 |  |  | M |
| Namazova-Baranova et al. (2020) | Case control | Russia | Assessed the sense of smell of children after COVID-19 infection | Non-hospitalised | Mean 11 (SD 3.5) | 39% | Ag | Mean 38 days | 61 | Unknown | 20 | Unknown | M |
| Erol et al. (2021) | Cohort | Turkey | Evaluated persisting COVID-19 related symptoms and to assessed cardiac findings to determine the impact of COVID-19 on children’s cardiovascular health | Mixed hospitalised / non-hospitalised | Median 9 (IQR 11-19) | 46% | Ag | 1+ months | 121 | 45 |  |  | H |
| Asadi-Pooya et al. (2021) | Cohort | Iran | Identified the prevalence and also the full spectrum of symptoms/complaints of children and adolescents who are suffering from long COVID | Hospitalised | Mean 12 (SD 3) | 52% | Ag | 3+ months | 58 | 26 |  |  | M |
| Malecki et al. (2021) | Case series | Poland | Described a group of pediatric patients with severe COVID-19, treated with convalescent plasma, to asses its effectiveness | Hospitalised | Median 12 (IQR 6–16) | 54% | Ag | 28 days | 13 | 4 |  |  | H |
| Prolonged symptoms case studies | | | | | | | | | | | | | |
| Ng (2020) | Case report | UK | Reported the prolonged dermatological manifestation 4 weeks following recovery from COVID-19 in a child | Non-hospitalised, secondary care | 12 | 0% | Ag | 7 weeks | 1 | 1 |  |  | L |
| Kahwagi et al. (2020) | Case report | Senegal | Reported the case of a 7-year-old female living in Senegal with encephalitis following infection with SARS-CoV-2. | Hospitalised | 7 | 100% | Ag | 2 months | 1 | 1 |  |  | H |
| Das (2021) | Case report | USA | Reported the case of a high school athlete with palpatations, myalgia, fatigue and dyspnea on exertion after SARS-CoV-2 infection | Non-hospitalised, secondary care | 16 | 100% | Ag | at least 2 months 6 days | 1 | 1 |  |  | M |
| Kumar et al. (2021) | Case report | USA | Reported a case of a 27-week-gestation extremely premature infant born to a mother with COVID-19 | Hospitalised | 27 week gestational age | 0% | Ag | 57 days | 1 | 1 |  |  | L |
| Sinaei et al. (2021) | Case report | Iran | Reported two cases of children diagnosed with post COVID reactive arthritis (only one had symptoms longer than 28 days) | Hospitalised | 8 | 100% | Ab | 5 weeks | 1 | 1 |  |  | M |
| Landzberg et al. (2021) | Case report | USA | Reported a case of Wernicke's encephalopathy in a girl with poor oral intake secondary to post-COVID-19 anosmia and dysgeusia | Hospitalised | 15 | 100% | Cl plus exposure | 3 months 1 week | 1 | 1 |  |  | M |
| Cecchini et al. (2022) | Case report | Italy | Reported a case of a 17-year-old female with anosmia after COVID-19, with partial recovery 15-months after the onset | Unclear | 17 | 100% | Ab | 15 months | 1 | 1 |  |  | L |
| Collins et al. (2022) | Case report | USA | Reported a case of severe costochondritis in a child who had previous COVID-19 | Hospitalised | 11 | 0% | Ag | 40 days | 1 | 1 |  |  | L |
| Ferreira et al. (2022) | Case report | Portugal | Reported a case of facial palsy in an otherwise healthy child folLing COVID-19 | Hospitalised | 11 | 0% | Ag | At least 6 months | 1 | 1 |  |  | L |
| Vu et al. (2021) | Case report | USA | Reported a case of a child with COVID-19 and Streptococcus pneumoniae bacterial coinfection | Hospitalised | 4 | 0% | Ag | 41 days | 1 | 1 |  |  | L |
| Thede (2021) | Case report | Germany | Discussed the signficance of long-Covid in children/adolescents and outlined possible (Chinese herbal) therapeutic concepts and concludes with 2 case studies (case study 1=adult) | Community recruitment | 12 | 100% | Ab | 6 weeks | 1 | 1 |  |  | M |
| Tomar et al. (2021) | Case report | India | Reported a case of a 13-year-old patient with acute post-infectious cerebellar ataxia following COVID-19 | Hospitalised | 13 | 0% | Ag | 42 days | 1 | 1 |  |  | M |
| Erdizci et al. (2021) | Case report | Turkey | Presented a case of simultaneous bilateral spontaneous pneumothorax with COVID-19 pneumonia | Hospitalised | 17 | 0% | Ag | 33 days | 1 | 1 |  |  | L |
| Clinical sequalae of acute COVID-19 | | | | | | | | | | | | | |
| Chevinsky et al. (2021) | Matched cohort | USA | Assessed the type, association and timing of post-COVID conditions in inpatient and outpatient settings | Mixed hospitalised / non-hospitalised | <16 years | Unknown | Ag | 4 months | 2673 | Unclear | 2673 | Unclear | M |
| LaRovere et al. (2021) | Case series | USA | Described the type and severity of neurologic involvement and documented hospital outcomes, using the Overcoming COVID-19 US public health surveillance registry of children and adolescents hospitalized with COVID-19–related complications | Hospitalised | Median 9 (IQR 2-15) | 46% | Ag | 1+ months | 365 | 7 |  |  | L |
| Thakur & Rai (2022) | Case series | India | Reported two cases of diabetic ketoacidosis / new onset type 1 diabetes with a recent history of COVID-19 | Not hospitalised | 2 and 5 | 0% | Cl plus Ab | 1+ months | 2 | 2 |  |  | M |
| Trieu et al. (2021) | Case series | USA | Illustrated an increase in the incidence of types 1 and 2 diabetes between April-November 2020 at a large tertiary care children's hospital and examined the characteristics and adverse outcomes in these children (some had simultaneous acute COVID-19 and new onset type 1 DM) | Hospitalised | Mean 11 | 70% | Ag | N/A | 10 | 10 |  |  | L |
| Boboc et al. (2021) | Observational retrospective cohort study | Romania | Reported a possible increase in the number of new type 1 diabetes diagnoses in children from Bucharest and the surrounding areas during the COVID-19-19 pandemic, compared to previous years, and evaluated predictors of diabetic ketoacidosis at disease onset (some had simultaneous acute COVID-19 and new onset type 1 DM) | Hospitalised | Mean 9 | 50% | Ag | N/A | 8 | 8 |  |  | M |
| Slae et al. (2021) | Case series | Israel (multi- national survey) | Described the extent to which COVID-19 may co-exist with acute pancreatitis in children (some developed pancreatic pseudocysts) | Hospitalised | Range 0-21 | 68% | Cl plus Ab or Ag | 1+ months | 22 | 3 |  |  | H |
| Chua et al. (2021) | Cross sectional study | Hong Kong | Compared the clinical characteristics and sources of infection among young people with COVID-19 during the 3 waves of outbreaks in Hong Kong 2020 (one subsequently developed haemolytic anaemia) | Community testing | Mean 10 (SD 5) | 45% | Ag | 3+ months | 397 | 1 |  |  | L |
| Clinical sequalae case studies | | | | | | | | | | | | | |
| Khalifa et al. (2020) | Case report | Pakistan | Reported a case of Guillain-Barre Syndrome associated with SARS-CoV-2 infection | Hospitalised | 11 | 0%% | Ag | 43 days | 1 | 1 |  |  | L |
| Nielsen-Saines et al. (2021) | Case report | USA | Reported a possible association between COVID-19 and type 1 diabetes in children | Hospitalised | 7 | 0% | Ag | 47 days | 1 | 1 |  |  | L |
| Akçay et al. (2021) | Case report | Turkey | Reported the clinical features of a child with axonal Guillain-Barre syndrome associated with SARS CoV-2 infection | Hospitalised | 6 | 0% | Ag | 60 days | 1 | 1 |  |  | M |
| Khera et al. (2021) | Case report | India | Reported the case of a 15-year-old girl with COVID-19 and acute focal deficit with altered sensorium due to massive right intracerebral hemorrhage following a hypertensive emergency and acute on chronic kidney disease | Hospitalised | 15 | 100% | Ag | 28+ days | 1 | 1 |  |  | L |
| Javed et al. (2021) | Case report | USA | Reported a case of a male adolescent who developed psychosis following COVID-19 | Not hospitalised | 17 | 0% | Ag | 11 months | 1 | 1 |  |  | M |
| Scala et al. (2021) | Case report | Italy | Reported the case of a child with serological evidence of SARS-CoV-2 infection whose onset was a massive right cerebral artery ischemia that led to a malignant cerebral infarction | Hospitalised | 11 | 0% | Cl plus Ab | 5+ weeks | 1 | 1 |  |  | L |
| Ordooei et al. (2021) | Case report | Iran | Reported the case of a child with new onset type 1 diabetes presenting with diabetic ketoacidosis, and concurrent COVID-19 | Hospitalised | 10 | 0% | Ab | 31 days | 1 | 1 |  |  | L |
| Mezzeoui et al. (2021) | Case report | Morocco | Reported the case of a patient diagnosed with post-Covid Guillan-Barre Syndrome | Hospitalised | 3 | 100% | Cl | 1 month | 1 | 1 |  |  | H |
| DeVette et al. (2021) | Case report | USA | Reported the case of a pediatric patient presenting with isolated choreiform movements who was ultimately diagnosed with acute rheumatic fever (ARF) and COVID-19. | Hospitalised | 8 | 100% | Ag | 1+ month | 1 | 1 |  |  | H |
| Shree et al. (2021) | Case report | India | Reported the case of an acute ischemic stroke in a young child associated with COVID-19 | Hospitalised | 1 | 100% | Ab | 4+ weeks | 1 | 1 |  |  | L |
| Persistent abnormal laboratory or radiological findings | | | | | | | | | | | | | |
| Denina et al. (2020) | Cohort | Italy | Evaluated the sequalae of COVID-19 in previously hospitalised children (persisting lung ultrasound changes and raised inflammatory markers) | Hospitalised | Median 8 (range 0-15) | 48% | Ag | Mean 35 days post discharge | 25 | 10 |  |  | M |
| Zhang et al. (2021) | Case series | China | Evaluated the pulmonary manifestations in and clinical characteristics of 14 paediatric patients with COVID‐19 (persisting chest CT abnormalities) | Hospitalised | Median 2 (IQR 0 – 5) | 71% | Cl or Ag | 30 days | 14 | 7 |  |  | M |
| Tang et al. (2021) | Cohort | China | Reported the clinical features and 1-month follow up observations for paediatric patients hospitalized with COVID-19 in Wuhan Women and Children's hospital (persisting chest CT abnormalities) | Hospitalised | Mean 5 (SD 4.3) | 33% | Ag | 1 month post discharge | 46 | 22 |  |  | H |
| Guemes-Villahoz et al. (2021) | Case control | Spain | Evaluated retinal vascular changes in children recovered from COVID-19 using OCTA and compared results with age-matched healthy children | Mixed hospitalised / non-hospitalised | Mean 12 (SD +-3) | 65% | Ag | Mean 38 days from diagnosis | 27 | Unknown | 45 | Unknown | L |
| Persistent abnormal laboratory or radiological findings case studies | | | | | | | | | | | | | |
| Shah & Carter (2020) | Case report | USA | Reported a case of nephotic syndrome in a child with COVID-19 infection | Non-hospitalised | 8 |  | Ag | 4+ weeks | 1 | 1 |  |  | H |
| Qiu et al. (2020) | Case report | China | Reported the clinical course and follow-up data of a critically ill infant with COVID-19 | Hospitalised | 8 months | 0% | Ag | 50+ days | 1 | 1 |  |  | H |
| Gerber et al. (2021) | Case report | USA | Reported the case of a 16-year-old asymptomatic male who presented with coronary artery dilation identifed on echo performed solely due to presence of COVID-19 antibodies | Non-hospitalised | 16 | 0% | Ab | 2+ months | 1 | 1 |  |  | H |
| Kossiva et al. (2021) | Case report | Greece | Reported the case of a 14-year old who presented with thrombocytopenia and leukopenia | Not hospitalised | 14 | 0% | Ab | 5 months | 1 | 1 |  |  | M |
| Manzo et al. (2021) | Case report | Italy | Reported a case of acute disseminated encephalomyelitis in a 6-year-old (underlying Fisher-Evans syndrome) with Sars-Cov2 infection | Hospitalised | 6 | 0% | Ab | 35 days | 1 | 1 |  |  | L |
| Khera et al. (2021) | Case report | India | Reported the case a child who presented with acute febrile illness followed by acute onset severe flaccid paralysis requiring prolonged intensive care unit stay and ventilator support. | Hospitalised | 11 | 100% | Ag | 6 weeks | 1 | 1 |  |  | L |
| Pre-existing conditions | | | | | | | | | | | | | |
| Berteloot et al. (2021) | Case series | France | Reported the occurrence of graft vascular anomalies in seven of nine children who received kidney transplants since the beginning of the COVID-19 pandemic | Hospitalised | Median 3 (range 3 - 17) | 11% | Ag or Ab | 3 months | 9 | 4 |  |  | L |
| Welzel et al. (2021) | Case series | Germany | Reported the course of COVID-19 in patients with IL-1-mediated and unclassified AID with immunosuppressive therapy | Non-hospitalised, secondary care | Mean 14 (range 12-15) | 67% | Cl or Ag | 40+ days | 3 | 3 |  |  | M |
| Barhoom et al. (2021) | Case series | Iran | Reported the clinical effects of COVID-19 in 4 children who were recipients of hematopoietic stem cell transplants | Hospitalised | Median 6 (range 3-10) | 25% | Ag | 1+ months | 4 | 2 |  |  | L |
| Kamdar et al. (2021) | Cohort | USA | Reported the characteristics and outcomes of COVID-19 in children with cancer or hematologic disorders | Mixed hospitalised / non-hospitalised | Median 10 (IQR 4-15) | 41% | Ag | 6 weeks | 109 | 1 |  |  | M |
| Conway et al. (2021) | Case series | USA | Described the patient population and infection related outcomes of COVID-19 on pediatric heart transplant candidates and recipients | Mixed hospitalised / non-hospitalised | Median 14 years (IQR 7-18) | 50% | Cl or Ag | 30 + days | 225 | 7 |  |  | M |
| Madhusoodhan et al. (2020) | Cohort | USA | Reported acute COVID-19 outcomes in childen with cancer | Mixed hospitalised / non-hospitalised | Median 13 (range 2-21) | 29% | Ag | 1+ months | 98 | Unknown |  |  | L |
| Hugle et al. (2021) | Case series | Germany | Reported 5 cases of patients with juvenile idiopathic arthritis in remission or long-term inactive disease on medication who developed flares of their disease in close temporal correlation with confirmed prior COVID-19 | Non-hospitalised | Median 15 (range 7-17) | 60% | Ag | 4+ weeks | 5 | 3 |  |  | L |
| Pre-existing conditions case studies | | | | | | | | | | | | | |
| Bush et al. (2020) | Case report | USA | Reported a case of COVID-19 in a paediatric renal transplant recipient | Hospitalised | 13 | 0% | Ag | 1+ month | 1 | 1 |  |  | L |
| DeVine et al. (2020) | Case report | USA | Reported a case of an immunocompromised child who was hospitalised several times with COVID-19 and safely treated with Remdesivir. | Hospitalised | 17 | 100% | Ag | 70+ days | 1 | 1 |  |  | L |
| Leclercq et al. (2020) | Case report | Switzerland | Reported the presentation of a newly diagnosed malignancy temporally associated with COVID-19 and a state of immunosupression | Non-hospitalised | 8 | 0% | Ab | 32 days | 1 | 1 |  |  | M |
| Ionescu et al. (2020) | Case report | Romania | Reported the case of a 13y-year-old old female patient with pre-existing renal failure secondary to PKD who developed acute pericarditis and bilateral pleurisy temporally associated with SARS-CoV-2 infection | Hospitalised | 13 | 100% | Ag | 2+ months | 1 | 1 |  |  | M |
| Aghaei Moghadam et al. (2021) | Case report | Iran | Reported the case of an 18-month old with a prolonged SARS-CoV-2 RNA shedding and chronic right atrial and superior vena cava (SVC) thrombosis | Hospitalised | 1 | 0% | Ag | 2+ months | 1 | 1 |  |  | L |
| Dongre et al. (2021) | Case report | India | Reported the case of a 5.5 year-old on maintenance chemotherapy for acute lymphoblastic leukaemia who subsequently developed immune thrombocytopenia secondary to SARS-CoV-2 infection | Hospitalised | 5 | 100% | Ag | 4+ months | 1 | 1 |  |  | L |
| Pereira et al. (2021) | Case report | UK | Reported the case of a boy with X-linked agammaglobulinemia who had mild acute COVID-19 but after recovering developed fevers and a raised erythrocyte sedimentation rate that persisted for several weeks | Hospitalised | 17 | 0% | Ag | 9 weeks | 1 | 1 |  |  | L |
| Truong et al. (2021) | Case report | USA | Reported prolonged SARS-CoV-2 RT-PCR positivity in two children and one young adult undergoing therapy for B-cell acute lymphoblastic leukemia (ALL). | Hospitalised | <5 | 0% | Ag | 6+ months | 1 | 1 |  |  | L |
| Koh et al. (2021) | Case report | USA | Reported the case of a child with sickle cell disease referred for lung transplant evaluation who presented with acute chest syndrome complicated by SARS-CoV-2 infection | Hospitalised | 11 | 0% | Nd | 5 months | 1 | 1 |  |  | M |
| * COVID-19 diagnosis method: Cl= Clinical, Ag=Antigen positive, Ab=Antibody positive, Cc=Close contact, SR=Self report, Nd=Not described |  |  |  |  |  |  |  |  |  | |  | |  |
| ** Risk of bias: H= High, M=Moderate, L=Low |  |  |  |  |  |  |  |  |  | |  | |  |

Supplementary material – S2 – PRISMA checklist

| **Section and Topic** | **Item #** | **Checklist item** | **Location where item is reported** |
| --- | --- | --- | --- |
| **TITLE** | | |  |
| Title | 1 | Identify the report as a systematic review. | Page 2 |
| **ABSTRACT** | | |  |
| Abstract | 2 | See the PRISMA 2020 for Abstracts checklist. (also attached) | Page 2 |
| **INTRODUCTION** | | |  |
| Rationale | 3 | Describe the rationale for the review in the context of existing knowledge. | Page 3-4 |
| Objectives | 4 | Provide an explicit statement of the objective(s) or question(s) the review addresses. | Page 4 |
| **METHODS** | | |  |
| Eligibility criteria | 5 | Specify the inclusion and exclusion criteria for the review and how studies were grouped for the syntheses. | Page 4 |
| Information sources | 6 | Specify all databases, registers, websites, organisations, reference lists and other sources searched or consulted to identify studies. Specify the date when each source was last searched or consulted. | Page 5 |
| Search strategy | 7 | Present the full search strategies for all databases, registers and websites, including any filters and limits used. | Supplementary data, Table S2 |
| Selection process | 8 | Specify the methods used to decide whether a study met the inclusion criteria of the review, including how many reviewers screened each record and each report retrieved, whether they worked independently, and if applicable, details of automation tools used in the process. | Page 5 |
| Data collection process | 9 | Specify the methods used to collect data from reports, including how many reviewers collected data from each report, whether they worked independently, any processes for obtaining or confirming data from study investigators, and if applicable, details of automation tools used in the process. | Page 5 |
| Data items | 10a | List and define all outcomes for which data were sought. Specify whether all results that were compatible with each outcome domain in each study were sought (e.g. for all measures, time points, analyses), and if not, the methods used to decide which results to collect. | Page 5 |
|  | 10b | List and define all other variables for which data were sought (e.g. participant and intervention characteristics, funding sources). Describe any assumptions made about any missing or unclear information. | Page 5 |
| Study risk of bias assessment | 11 | Specify the methods used to assess risk of bias in the included studies, including details of the tool(s) used, how many reviewers assessed each study and whether they worked independently, and if applicable, details of automation tools used in the process. | Page 5 |
| Effect measures | 12 | Specify for each outcome the effect measure(s) (e.g. risk ratio, mean difference) used in the synthesis or presentation of results. Same as effect estimate? | Page 6 |
| Synthesis methods | 13a | Describe the processes used to decide which studies were eligible for each synthesis (e.g. tabulating the study intervention characteristics and comparing against the planned groups for each synthesis (item #5)). | Page 5-6 |
|  | 13b | Describe any methods required to prepare the data for presentation or synthesis, such as handling of missing summary statistics, or data conversions. | N/A |
|  | 13c | Describe any methods used to tabulate or visually display results of individual studies and syntheses. | Page 6 |
|  | 13d | Describe any methods used to synthesize results and provide a rationale for the choice(s). If meta-analysis was performed, describe the model(s), method(s) to identify the presence and extent of statistical heterogeneity, and software package(s) used. | Page 6 |
|  | 13e | Describe any methods used to explore possible causes of heterogeneity among study results (e.g. subgroup analysis, meta-regression). | Page 6 |
|  | 13f | Describe any sensitivity analyses conducted to assess robustness of the synthesized results. | Page 6 |
| Reporting bias assessment | 14 | Describe any methods used to assess risk of bias due to missing results in a synthesis (arising from reporting biases). | Page 6 |
| Certainty assessment | 15 | Describe any methods used to assess certainty (or confidence) in the body of evidence for an outcome. | Page 6 |
| **RESULTS** | | |  |
| Study selection | 16a | Describe the results of the search and selection process, from the number of records identified in the search to the number of studies included in the review, ideally using a flow diagram. | Page 6 and FIgure 1 |
|  | 16b | Cite studies that might appear to meet the inclusion criteria, but which were excluded, and explain why they were excluded. | Figure 1 |
| Study characteristics | 17 | Cite each included study and present its characteristics. | Supplementary material – Table 1 |
| Risk of bias in studies | 18 | Present assessments of risk of bias for each included study. | Supplementary material – Table 1 |
| Results of individual studies | 19 | For all outcomes, present, for each study: (a) summary statistics for each group (where appropriate) and (b) an effect estimate and its precision (e.g. confidence/credible interval), ideally using structured tables or plots. | Pages 6 and Figures 3-5 |
| Results of syntheses | 20a | For each synthesis, briefly summarise the characteristics and risk of bias among contributing studies. | Pages 6-9 |
|  | 20b | Present results of all statistical syntheses conducted. If meta-analysis was done, present for each the summary estimate and its precision (e.g. confidence/credible interval) and measures of statistical heterogeneity. If comparing groups, describe the direction of the effect. | Pages 6-9 |
|  | 20c | Present results of all investigations of possible causes of heterogeneity among study results. | Pages 7-8 |
|  | 20d | Present results of all sensitivity analyses conducted to assess the robustness of the synthesized results. | Pages 6-9 |
| Reporting biases | 21 | Present assessments of risk of bias due to missing results (arising from reporting biases) for each synthesis assessed. | Page 7 |
| Certainty of evidence | 22 | Present assessments of certainty (or confidence) in the body of evidence for each outcome assessed. | Pages 6-9 |
| **DISCUSSION** | | |  |
| Discussion | 23a | Provide a general interpretation of the results in the context of other evidence. | Pages 9-11 |
|  | 23b | Discuss any limitations of the evidence included in the review. | Page 11 |
|  | 23c | Discuss any limitations of the review processes used. | Page 11 |
|  | 23d | Discuss implications of the results for practice, policy, and future research. | Page 11 |
| **OTHER INFORMATION** | | |  |
| Registration and protocol | 24a | Provide registration information for the review, including register name and registration number, or state that the review was not registered. | Page 4 |
|  | 24b | Indicate where the review protocol can be accessed, or state that a protocol was not prepared. | Page 4 |
|  | 24c | Describe and explain any amendments to information provided at registration or in the protocol. | N/A |
| Support | 25 | Describe sources of financial or non-financial support for the review, and the role of the funders or sponsors in the review. | Page 6 |
| Competing interests | 26 | Declare any competing interests of review authors. | Page 11 |
| Availability of data, code and other materials | 27 | Report which of the following are publicly available and where they can be found: template data collection forms; data extracted from included studies; data used for all analyses; analytic code; any other materials used in the review. | Page 11 |

*From:* Page MJ, McKenzie JE, Bossuyt PM, Boutron I, Hoffmann TC, Mulrow CD, et al. The PRISMA 2020 statement: an updated guideline for reporting systematic reviews. BMJ 2021;372:n71. doi: 10.1136/bmj.n71

For more information, visit: <http://www.prisma-statement.org/>

| **Section and Topic** | **Item #** | **Checklist item** | **Reported (Yes/No)** |
| --- | --- | --- | --- |
| **TITLE** | | |  |
| Title | 1 | Identify the report as a systematic review. | Y |
| **BACKGROUND** | | |  |
| Objectives | 2 | Provide an explicit statement of the main objective(s) or question(s) the review addresses. | Y |
| **METHODS** | | |  |
| Eligibility criteria | 3 | Specify the inclusion and exclusion criteria for the review. | Y |
| Information sources | 4 | Specify the information sources (e.g. databases, registers) used to identify studies and the date when each was last searched. | Y |
| Risk of bias | 5 | Specify the methods used to assess risk of bias in the included studies. | Y |
| Synthesis of results | 6 | Specify the methods used to present and synthesise results. | Y |
| **RESULTS** | | |  |
| Included studies | 7 | Give the total number of included studies and participants and summarise relevant characteristics of studies. | Y |
| Synthesis of results | 8 | Present results for main outcomes, preferably indicating the number of included studies and participants for each. If meta-analysis was done, report the summary estimate and confidence/credible interval. If comparing groups, indicate the direction of the effect (i.e. which group is favoured). | Y |
| **DISCUSSION** | | |  |
| Limitations of evidence | 9 | Provide a brief summary of the limitations of the evidence included in the review (e.g. study risk of bias, inconsistency and imprecision). | Y |
| Interpretation | 10 | Provide a general interpretation of the results and important implications. | Y |
| **OTHER** | | |  |
| Funding | 11 | Specify the primary source of funding for the review. | Y |
| Registration | 12 | Provide the register name and registration number. | Y |

*From:* Page MJ, McKenzie JE, Bossuyt PM, Boutron I, Hoffmann TC, Mulrow CD, et al. The PRISMA 2020 statement: an updated guideline for reporting systematic reviews. BMJ 2021;372:n71. doi: 10.1136/bmj.n71

For more information, visit: <http://www.prisma-statement.org/>

Supplementary material – S3 – Search strategies

SPLaT-19 study: Initial search 05.01.2021

### Search record

| Date | | **Database/**  **Info source** | **Search From** | | | **Terms** | **Results** |
| --- | --- | --- | --- | --- | --- | --- | --- |
| 05.01.2021 | | Medline (Ovid) | 01.12.2019 | | | See below | 481 |
| 05.01.2021 | | EMBASE (Ovid) | 01.12.2019 | | | See below | 301 |
| 05.01.2021 | | AMED (Ovid) | Inception | | | See below | 1 |
| 05.01.2021 | | HMIC (DoH & Kings fund) (Ovid) | Inception | | | See below | 3 |
| 05.01.2021 | | CINAHLPlus (EBSCO) | December 2019 | | | See below | 149 |
| 05.01.2021 | | APA PsycINFO (EBSCO) | December 2019 | | | See below | 53 |
| 05.01.2021 | | Web of Science (SCI-EXPANDED, SSCI) | 01.12.2019 | | | See below | 260 |
| 05.01.2021 | | ASSIA (ProQuest) | 01.12.2019 | | | See below | 63 |
| 05.01.2021 | | WHO Covid-19: Global literature on coronavirus disease | 01.12.2019 | | | See below | 450 |
| 05.01.2021 | | Cochrane Covid-19 study register | 01.12.2019 | | | See below | 610* |
| 05.01.2021 | | ProQuest Coronavirus research database | 01.12.2019 | | | See below | 514 |
| 05.01.2021 | | NDLTD | 2019 | | See below | | 24 |
| 05.01.2021 | | OpenGrey | 2019 | | See below | | 0 |
|  | |  |  | **Total number of records =** | | | **2909** |
|  |  | |  | **Records after de-duplication =** | | | **1914** |

*this excludes records from trials registers (clinicaltrials.gov & ICTRP) **n**=237 but includes multiple citations

### Search strategies

**For Ovid:** The following table is an explanation of the symbols used in the search strategy below.

/	indicates an index term (MeSH/EMTREE heading)

exp	before an index term indicates that all subheadings were selected

.ti,ab,kf.	indicates a search for a term in title/abstract/word(s) in keyword [MEDLINE]

.rx,px,ox.	indicates a search for a term in rare disease-, protocol-, organism-supplementary concept word [MEDLINE]

.os.	indicates a search for a term in organism supplementary concept [MEDLINE]

*[*n*]	at the end of a term indicates that this term has been truncated [by *n* character(s)].

adj	indicates a search for two terms where they appear adjacent to each another

adj*n*	indicates a search for two terms where they appear within *n* words of each another [OVID]

Medline (OvidSP) 05.01.2021 - Ovid MEDLINE(R) ALL 1946 to January 04, 2021

| 1 | exp Child/ | 1936558 |
| --- | --- | --- |
| 2 | exp infant/ | 1152655 |
| 3 | Adolescent/ | 2058510 |
| 4 | Pediatrics/ | 54654 |
| 5 | (paediatri* or pediatri*).ti,ab,kf. | 382927 |
| 6 | child*.ti,ab,kf. | 1468417 |
| 7 | toddler*.ti,ab,kf. | 11716 |
| 8 | infant*.ti,ab,kf. | 457416 |
| 9 | (baby or babies).ti,ab,kf. | 72291 |
| 10 | neonate*.ti,ab,kf. | 94316 |
| 11 | (newborn* or new born*).ti,ab,kf. | 183536 |
| 12 | girl*.ti,ab,kf. | 152467 |
| 13 | boy*.ti,ab,kf. | 157497 |
| 14 | preschool*.ti,ab,kf. | 30241 |
| 15 | school*.ti,ab,kf. | 294228 |
| 16 | adolescen*.ti,ab,kf. | 309800 |
| 17 | teen*.ti,ab,kf. | 31512 |
| 18 | youth*.ti,ab,kf. | 84290 |
| 19 | juvenile*.ti,ab,kf. | 84905 |
| 20 | (young adj (person or people)).ti,ab,kf. | 29402 |
| 21 | kindergarten*.ti,ab,kf. | 6795 |
| 22 | or/1-21 [CYP terms ie 0-17 years] | 4436343 |
| 23 | ((longterm or long) adj4 (covid* or corona or coronavirus*)).ti,ab,kf. | 437 |
| 24 | (chronic adj3 (covid* or corona or coronavirus*)).ti,ab,kf. | 110 |
| 25 | (prolong* adj3 (covid* or corona or coronavirus*)).ti,ab,kf. | 84 |
| 26 | (persist* adj3 (covid* or corona or coronavirus*)).ti,ab,kf. | 131 |
| 27 | (sustain* adj3 (covid* or corona or coronavirus*)).ti,ab,kf. | 68 |
| 28 | (history adj3 (covid* or corona or coronavirus*)).ti,ab,kf. | 192 |
| 29 | ((post or postacute or postvir*) adj4 (covid* or corona or coronavirus*)).ti,ab,kf. | 915 |
| 30 | or/23-29 [Long-COVID specific terms] | 1884 |
| 31 | exp coronavirus/ | 45361 |
| 32 | exp coronavirus infections/ | 49604 |
| 33 | ((novel or new or nouveau) adj2 (CoV or Pandemi*2)).mp. | 1937 |
| 34 | (coronavir* or corona vir* or OC43 or NL63 or 229E or HKU1 or HCoV* or NCoV* or covid* or sarscov*).mp. | 104626 |
| 35 | betacorona*.mp. | 33603 |
| 36 | exp pneumonia/ and (wuhan or hubei or huanan).mp. | 2258 |
| 37 | ((pneumonia or SARS or severe acute respiratory) and (wuhan or hubei or huanan)).mp. | 3789 |
| 38 | ((wuhan or hubei or huanan) adj virus).mp. | 16 |
| 39 | (2019nCoV* or CoV2 or CoV 2).mp. | 32638 |
| 40 | (COVID-19 or severe acute respiratory syndrome coronavirus 2).os. [supplementary concepts] | 33013 |
| 41 | or/31-40 [COVID-19 terms] | 111745 |
| 42 | ((longterm or long term) adj3 (complication* or infect* or symptom* or syndrome* or consequence* or outcome* or impact* or suffer* or effect* or debilit*)).ti,ab,kf. | 200541 |
| 43 | ((chronic or persist* or prolong* or sustain* or continu*) adj3 (complication* or infect* or symptom* or syndrome* or consequence* or outcome* or impact* or suffer* or effect* or debilit*)).ti,ab,kf. | 287295 |
| 44 | ((post or postvir* or postacute) adj4 (complication* or infect* or symptom* or syndrome* or consequence* or outcome* or impact* or suffer* or effect* or debilit*)).ti,ab,kf. | 92464 |
| 45 | ((after or following) adj infect*).ti,ab,kf. | 32723 |
| 46 | (long haul* or longhaul*).ti,ab,kf. | 864 |
| 47 | ((prolong* or extend* or lengthen* or postpone* or delay*) adj3 (recovery or convalescen* or rehab*)).ti,ab,kf. | 10190 |
| 48 | sequela*.ti,ab,kf. | 70637 |
| 49 | or/42-48 [long-term terms] | 663886 |
| 50 | 41 and 49 [COVID and long-term combined] | 3824 |
| 51 | 30 or 50 [ALL terms re Long-COVID] | 5316 |
| 52 | 22 and 51 [Long COVID and CYP] | 582 |
| 53 | exp animals/ not humans/ | 4772383 |
| 54 | 52 not 53 | 562 |
| 55 | limit 54 to ed=20191201-20210105 [entry date: when processing of record ends] | 264 |
| 56 | limit 54 to dt=20191201-20210105 [Create date - date added to PubMed] | 476 |
| 57 | 55 or 56 | 481 |

EMBASE (OvidSP) 05.01.2021 – EMBASE 1974 to 2020 January 04 (note whilst 2020 is stated, results run to 2021)

| 1 | exp Child/ | 2689428 |
| --- | --- | --- |
| 2 | exp pediatrics/ | 110141 |
| 3 | juvenile/ | 46031 |
| 4 | exp Adolescent/ | 1561994 |
| 5 | (paediatri* or pediatri*).ti,ab,kw. | 599027 |
| 6 | child*.ti,ab,kw. | 1807011 |
| 7 | toddler*.ti,ab,kw. | 15320 |
| 8 | infant*.ti,ab,kw. | 483128 |
| 9 | (baby or babies).ti,ab,kw. | 100898 |
| 10 | neonate*.ti,ab,kw. | 128692 |
| 11 | (newborn* or new born*).ti,ab,kw. | 204759 |
| 12 | girl*.ti,ab,kw. | 203383 |
| 13 | boy*.ti,ab,kw. | 210734 |
| 14 | school*.ti,ab,kw. | 358249 |
| 15 | preschool*.ti,ab,kw. | 36164 |
| 16 | adolescen*.ti,ab,kw. | 389485 |
| 17 | teen*.ti,ab,kw. | 43353 |
| 18 | youth*.ti,ab,kw. | 98780 |
| 19 | juvenile*.ti,ab,kw. | 102002 |
| 20 | kindergarten*.ti,ab,kw. | 7717 |
| 21 | (young adj (person or people)).ti,ab,kw. | 41104 |
| 22 | or/1-21 [CYP terms ie 0-17 years] | 4467167 |
| 23 | ((longterm or long) adj4 (covid* or corona or coronavirus*)).ti,ab,kw. | 416 |
| 24 | (chronic adj3 (covid* or corona or coronavirus*)).ti,ab,kw. | 121 |
| 25 | (prolong* adj3 (covid* or corona or coronavirus*)).ti,ab,kw. | 93 |
| 26 | (persist* adj3 (covid* or corona or coronavirus*)).ti,ab,kw. | 130 |
| 27 | (sustain* adj3 (covid* or corona or coronavirus*)).ti,ab,kw. | 59 |
| 28 | (history adj3 (covid* or corona or coronavirus*)).ti,ab,kw. | 199 |
| 29 | ((post or postacute or postvir*) adj4 (covid* or corona or coronavirus*)).ti,ab,kw. | 842 |
| 30 | or/23-29 [Long-COVID specific terms] | 1809 |
| 31 | exp Coronavirinae/ | 22723 |
| 32 | exp coronavirus infections/ | 24359 |
| 33 | ((novel or new or nouveau) adj2 (CoV or Pandemi*2)).mp. | 2097 |
| 34 | (coronavir* or corona vir* or OC43 or NL63 or 229E or HKU1 or HCoV* or NCoV* or covid* or sarscov*).mp. | 115424 |
| 35 | betacorona*.mp. | 8591 |
| 36 | exp pneumonia/ and (wuhan or hubei or huanan).mp. | 1293 |
| 37 | ((pneumonia or SARS or severe acute respiratory) and (wuhan or hubei or huanan)).mp. | 3593 |
| 38 | ((wuhan or hubei or huanan) adj virus).mp. | 11 |
| 39 | (2019nCoV* or CoV2 or CoV 2).mp. | 27976 |
| 40 | or/31-39 [COVID-19 terms] | 123576 |
| 41 | ((longterm or long term) adj3 (complication* or infect* or symptom* or syndrome* or consequence* or outcome* or impact* or suffer* or effect* or debilit*)).ti,ab,kw. | 297223 |
| 42 | ((chronic or persist* or prolong* or sustain* or continu*) adj3 (complication* or infect* or symptom* or syndrome* or consequence* or outcome* or impact* or suffer* or effect* or debilit*)).ti,ab,kw. | 397433 |
| 43 | ((post or postvir* or postacute) adj4 (complication* or infect* or symptom* or syndrome* or consequence* or outcome* or impact* or suffer* or effect* or debilit*)).ti,ab,kw. | 159040 |
| 44 | ((after or following) adj infect*).ti,ab,kw. | 37457 |
| 45 | (long haul* or longhaul*).ti,ab,kw. | 952 |
| 46 | sequela*.ti,ab,kw. | 88109 |
| 47 | ((prolong* or extend* or lengthen* or postpone* or delay*) adj3 (recovery or convalescen* or rehab*)).ti,ab,kw. | 14237 |
| 48 | or/41-46 [long-term terms] | 933725 |
| 49 | 40 and 48 [COVID and long-term combined] | 4347 |
| 50 | 30 or 49 [ALL terms re Long-COVID] | 5777 |
| 51 | 22 and 50 [Long COVID and CYP] | 653 |
| 52 | exp animal/ not human/ | 4898633 |
| 53 | 51 not 52 | 614 |
| 54 | limit 53 to dc=20191201-20210105 [date created - date of last activity on citation before delivered to ovid i.e. original file created] | 424 |
| 55 | limit 53 to dd=20191201-20210105 [date created for delivery state = "new" original info delivered to ovid] | 120 |
| 56 | 54 or 55 | 424 |
| 57 | limit 56 to embase | 301 |

AMED (OvidSP) 05.01.2021 – AMED (Allied and Complementary Medicine) 1985 to November 2020

| 1 | exp Child/ | 18423 |
| --- | --- | --- |
| 2 | exp infant/ | 2193 |
| 3 | exp Adolescent/ | 6107 |
| 4 | Pediatrics/ | 679 |
| 5 | (paediatri* or pediatri*).ti,ab. | 3405 |
| 6 | child*.ti,ab. | 19334 |
| 7 | toddler*.ti,ab. | 206 |
| 8 | infant*.ti,ab. | 1823 |
| 9 | (baby or babies).ti,ab. | 446 |
| 10 | neonate*.ti,ab. | 144 |
| 11 | (newborn* or new born*).ti,ab. | 244 |
| 12 | girl*.ti,ab. | 1342 |
| 13 | boy*.ti,ab. | 1501 |
| 14 | preschool*.ti,ab. | 686 |
| 15 | school*.ti,ab. | 5338 |
| 16 | adolescen*.ti,ab. | 4168 |
| 17 | teen*.ti,ab. | 295 |
| 18 | youth*.ti,ab. | 1323 |
| 19 | juvenile*.ti,ab. | 391 |
| 20 | (young adj (person or people)).ti,ab. | 723 |
| 21 | kindergarten*.ti,ab. | 144 |
| 22 | or/1-21 [CYP terms ie 0-17 years] | 34063 |
| 23 | ((longterm or long) adj4 (covid* or corona or coronavirus*)).ti,ab. | 1 |
| 24 | (chronic adj3 (covid* or corona or coronavirus*)).ti,ab. | 0 |
| 25 | (prolong* adj3 (covid* or corona or coronavirus*)).ti,ab. | 0 |
| 26 | (persist* adj3 (covid* or corona or coronavirus*)).ti,ab. | 0 |
| 27 | (sustain* adj3 (covid* or corona or coronavirus*)).ti,ab. | 0 |
| 28 | (history adj3 (covid* or corona or coronavirus*)).ti,ab. | 0 |
| 29 | ((post or postacute or postvir*) adj4 (covid* or corona or coronavirus*)).ti,ab. | 0 |
| 30 | or/23-29 [Long-COVID specific terms] | 1 |
| 31 | severe acute respiratory syndrome/ | 15 |
| 32 | ((novel or new or nouveau) adj2 (CoV or Pandemi*2)).mp. | 1 |
| 33 | (coronavir* or corona vir* or OC43 or NL63 or 229E or HKU1 or HCoV* or NCoV* or covid* or sarscov*).mp. | 20 |
| 34 | betacorona*.mp. | 0 |
| 35 | exp pneumonia/ and (wuhan or hubei or huanan).mp. | 0 |
| 36 | ((pneumonia or SARS or severe acute respiratory) and (wuhan or hubei or huanan)).mp. | 0 |
| 37 | ((wuhan or hubei or huanan) adj virus).mp. | 0 |
| 38 | (2019nCoV* or CoV2 or CoV 2).mp. | 1 |
| 39 | or/31-38 [COVID-19 terms] | 34 |
| 40 | ((longterm or long term) adj3 (complication* or infect* or symptom* or syndrome* or consequence* or outcome* or impact* or suffer* or effect* or debilit*)).ti,ab. | 2606 |
| 41 | ((chronic or persist* or prolong* or sustain* or continu*) adj3 (complication* or infect* or symptom* or syndrome* or consequence* or outcome* or impact* or suffer* or effect* or debilit*)).ti,ab. | 6077 |
| 42 | ((post or postvir* or postacute) adj4 (complication* or infect* or symptom* or syndrome* or consequence* or outcome* or impact* or suffer* or effect* or debilit*)).ti,ab. | 1648 |
| 43 | ((after or following) adj infect*).ti,ab. | 4978 |
| 44 | (long haul* or longhaul*).ti,ab. | 8 |
| 45 | sequela*.ti,ab. | 749 |
| 46 | ((prolong* or extend* or lengthen* or postpone* or delay*) adj3 (recovery or convalescen* or rehab*)).ti,ab. | 357 |
| 47 | or/41-46 [long-term terms] | 12982 |
| 48 | 39 and 47 [COVID and long-term combined] | 14 |
| 49 | 30 or 48 [ALL terms re Long-COVID] | 15 |
| 50 | 22 and 49 [Long COVID and CYP] | 1 |

HMIC (OvidSP) 05.01.2021 – HMIC Health Management Information Consortium 1979 to November 2020

| [# ▲](https://ovidsp.dc1.ovid.com/ovid-b/ovidweb.cgi?&S=CKELFPGMPHACLABAKPAKOEDKBIDHAA00&Sort+Sets=descending) | **Searches** | **Results** |
| --- | --- | --- |
| 1 | exp Children/ | 20142 |
| 2 | exp young people/ | 10859 |
| 3 | exp paediatrics/ | 619 |
| 4 | (paediatri* or pediatri*).ti,ab. | 2608 |
| 5 | child*.ti,ab. | 30922 |
| 6 | toddler*.ti,ab. | 124 |
| 7 | infant*.ti,ab. | 2885 |
| 8 | (baby or babies).ti,ab. | 2479 |
| 9 | neonate*.ti,ab. | 240 |
| 10 | (newborn* or new born*).ti,ab. | 609 |
| 11 | girl*.ti,ab. | 1363 |
| 12 | boy*.ti,ab. | 1212 |
| 13 | preschool*.ti,ab. | 327 |
| 14 | school*.ti,ab. | 8715 |
| 15 | adolescen*.ti,ab. | 4014 |
| 16 | teen*.ti,ab. | 1592 |
| 17 | youth*.ti,ab. | 2136 |
| 18 | juvenile*.ti,ab. | 560 |
| 19 | (young adj (person or people)).ti,ab. | 5490 |
| 20 | kindergarten*.ti,ab. | 59 |
| 21 | or/1-20 [CYP terms ie 0-17 years] | 50352 |
| 22 | ((longterm or long) adj4 (covid* or corona or coronavirus*)).ti,ab. | 9 |
| 23 | (chronic adj3 (covid* or corona or coronavirus*)).ti,ab. | 0 |
| 24 | (prolong* adj3 (covid* or corona or coronavirus*)).ti,ab. | 0 |
| 25 | (persist* adj3 (covid* or corona or coronavirus*)).ti,ab. | 0 |
| 26 | (sustain* adj3 (covid* or corona or coronavirus)).ti,ab. | 3 |
| 27 | (history adj3 (covid* or corona or coronavirus)).ti,ab. | 0 |
| 28 | ((post or postacute or postvir*) adj4 (covid* or corona or coronavirus*)).ti,ab. | 18 |
| 29 | or/22-28 [Long-COVID specific terms] | 30 |
| 30 | coronaviruses/ | 331 |
| 31 | severe acute respiratory syndrome/ | 161 |
| 32 | ((novel or new or nouveau) adj2 (CoV or Pandemi*2)).mp. | 39 |
| 33 | (coronavir* or corona vir* or OC43 or NL63 or 229E or HKU1 or HCoV* or NCoV* or covid* or sarscov*).mp. | 944 |
| 34 | betacorona*.mp. | 0 |
| 35 | exp pneumonia/ and (wuhan or hubei or huanan).mp. | 0 |
| 36 | ((pneumonia or SARS or severe acute respiratory) and (wuhan or hubei or huanan)).mp. | 0 |
| 37 | ((wuhan or hubei or huanan) adj virus).mp. | 0 |
| 38 | (2019nCoV* or CoV2 or CoV 2).mp. | 14 |
| 39 | or/30-38 [COVID-19 terms] | 1123 |
| 40 | ((chronic or persist* or prolong* or sustain* or continu*) adj3 (complication* or infect* or symptom* or syndrome* or consequence* or outcome* or impact* or suffer* or effect* or debilit*)).ti,ab. | 1874 |
| 41 | ((longterm or long term) adj3 (complication* or infect* or symptom* or syndrome* or consequence* or outcome* or impact* or suffer* or effect* or debilit*)).ti,ab. | 1418 |
| 42 | ((post or postvir* or postacute) adj4 (complication* or infect* or symptom* or syndrome* or consequence* or outcome* or impact* or suffer* or effect* or debilit*)).ti,ab. | 456 |
| 43 | ((after or following) adj infect*).ti,ab. | 24 |
| 44 | (long haul* or longhaul*).ti,ab. | 15 |
| 45 | sequela*.ti,ab. | 188 |
| 46 | ((prolong* or extend* or lengthen* or postpone* or delay*) adj3 (recovery or convalescen* or rehab*)).ti,ab. | 46 |
| 47 | or/40-44 [long-term terms] | 3676 |
| 48 | 39 and 47 [COVID and long-term combined] | 15 |
| 49 | 29 or 48 [ALL terms re Long-COVID] | 43 |
| 50 | 21 and 49 [Long COVID and CYP] | 3 |

CINAHLPlus (EBSCO) 05.01.2021

| S1 | (MH "Child+") | 684,653 |
| --- | --- | --- |
| S2 | (MH "Adolescence") | 540,362 |
| S3 | (MH "Pediatrics+") | 21,607 |
| S4 | TI (paediatri* or pediatri*) OR AB (paediatri* or pediatri*) | 141,541 |
| S5 | TI child* OR AB child* | 501,665 |
| S6 | TI toddler* OR AB toddler* | 6,754 |
| S7 | TI infant* OR AB infant* | 109,518 |
| S8 | TI ( baby or babies ) OR AB ( baby or babies ) | 34,005 |
| S9 | TI neonate* OR AB neonate* | 22,818 |
| S10 | TI ( newborn* or (new W0 born*) ) OR AB ( newborn* or (new W0 born*) ) | 31,969 |
| S11 | TI girl* OR AB girl* | 43,982 |
| S12 | TI boy* OR AB boy* | 42,913 |
| S13 | TI preschool* OR AB preschool* | 13,759 |
| S14 | TI school* OR AB school* | 146,758 |
| S15 | TI adolescen* OR AB adolescen* | 138,025 |
| S16 | TI teen* OR AB teen* | 19,345 |
| S17 | TI youth* OR AB youth* | 51,408 |
| S18 | TI juvenile* OR AB juvenile* | 10,333 |
| S19 | TI ( young W0 (person* or people) ) OR AB ( young W0 (person* or people) ) | 21,136 |
| S20 | TI kindergarten* OR AB kindergarten* | 2,941 |
| S21 | S1 OR S2 OR S3 OR S4 OR S5 OR S6 OR S7 OR S8 OR S9 OR S10 OR S11 OR S12 OR S13 OR S14 OR S15 OR S16 OR S17 OR S18 OR S19 OR S20 | 1,318,456 |
| S22 | TI (sustain* N3 (covid* OR corona OR coronavirus*)) OR AB (sustain* N3 (covid* OR corona OR coronavirus*)) | 47 |
| S23 | TI ( ("long term" or longterm) N4 (covid* or corona or coronavirus*) ) OR AB ( ("long term" or longterm) N4 (covid* or corona or coronavirus*) ) | 203 |
| S24 | TI ( chronic N2 (covid* or corona or coronavirus*) ) OR AB ( chronic N2 (covid* or corona or coronavirus*) ) | 43 |
| S25 | TI ( prolong* N2 (covid* or corona or coronavirus*) ) OR AB ( prolong* N2 (covid* or corona or coronavirus*) ) | 27 |
| S26 | TI ((persist* or history) N2 (covid* or corona or coronavirus*) ) OR AB ((persist* or history) N2 (covid* or corona or coronavirus*) ) | 86 |
| S27 | TI ( (post or postacute or postvir*) N2 (covid* or corona or coronavirus*) ) OR AB ( (post or postacute or postvir*) N2 (covid* or corona or coronavirus*) ) | 216 |
| S28 | S22 OR S23 OR S24 OR S25 OR S26 OR S27 | 607 |
| S29 | (MH "Coronavirus+") | 13,222 |
| S30 | (MH "Coronavirus Infections+") | 23,416 |
| S31 | ((novel or new or nouveau) N1 (CoV or Pandemi*) | 384 |
| S32 | coronavir* or (corona W0 vir*) or OC43 or NL63 or 229E or HKU1 or HCoV* or NCoV* or covid* or (sars W0 cov*) or sarscov* | 36,036 |
| S33 | betacorona* | 54 |
| S34 | (wuhan or hubei or huanan) W0 virus | 4 |
| S35 | 2019nCoV* or CoV2 or "CoV 2" or CoV-2 | 129 |
| S36 | (MH "Pneumonia+") | 31,512 |
| S37 | wuhan or hubei or huanan | 1,624 |
| S38 | S29 OR S30 OR S31 OR S32 OR S33 OR S34 OR S35 OR (S36 AND S37) | 38,020 |
| S39 | TI ((chronic or persist* or longterm or "long term" or prolong* OR sustain* OR continu*) N2 (complication* or infect* or symptom* or syndrome* or consequence* or outcome* or impact* or effect* or debilit*)) OR AB ((chronic or persist* or longterm or "long term" or prolong* OR sustain* OR continu*) N2 (complication* or infect* or symptom* or syndrome* or consequence* or outcome* or impact* or effect* or debilit*)) | 104,627 |
| S40 | TI ( (post or postvir*) N3 (complication* or infect* or symptom* or syndrome* or consequence* or outcome* or impact* OR suffer* or effect* or debilit*) ) OR AB ( (post or postvir*) N3 (complication* or infect* or symptom* or syndrome* or consequence* or outcome* or impact* OR suffer* or effect* or debilit*) ) | 20,923 |
| S41 | TI ( (long W0 haul*) or longhaul* or sequela*) OR AB ( (long W0 haul*) or longhaul* or sequela*) | 14,307 |
| S42 | TI ((after or following) W0 infect*) OR AB ((after or following) W0 infect*) | 1,332 |
| S43 | TI ( (prolong* or extend* or lengthen* or postpone* or delay*) N2 (recovery or convalescen* or rehab*)) OR AB ( (prolong* or extend* or lengthen* or postpone* or delay*) N2 (recovery or convalescen* or rehab*)) | 2,477 |
| S44 | S39 OR S40 OR S41 OR S42 OR S43 | 139,738 |
| S45 | S38 AND S44 | 694 |
| S46 | S28 OR S45 | 1,173 |
| S47 | S21 AND S46 | 163 |
| S48 | S21 AND S46  **Limiters** - Published Date: 20191201-20210131 | 149 |

APA PsycINFO (EBSCO) 05.01.2021

| S1 | DE "Child Characteristics" OR DE "Child Self Care" OR DE "Child Psychotherapy" OR DE "Child Psychopathology" | 12,837 |
| --- | --- | --- |
| S2 | DE "Early Adolescence" OR DE "Adolescent Characteristics" OR DE "Adolescent Psychopathology" OR DE "Adolescent Psychology" OR DE "Adolescent Health" OR DE "Adolescent Attitudes" OR DE "Adolescent Psychotherapy" OR DE "Adolescent Psychiatry" OR DE "Adolescent Behavior" | 57,195 |
| S3 | DE "Pediatrics" OR DE "Kindergarten Students" OR DE "Primary School Students" OR DE "Preschool Students" OR DE "Nursery School Students" OR DE "Middle School Students" OR DE "Junior High School Students" OR DE "Intermediate School Students" OR DE "High School Students" OR DE "Elementary School Students" OR DE "Community College Students" OR DE "College Students" OR DE "Vocational School Students" OR DE "Special Education Students" | 214,747 |
| S4 | TI (paediatri* or pediatri*) OR AB (paediatri* or pediatri*) | 37,539 |
| S5 | TI child* OR AB child* | 701,233 |
| S6 | TI toddler* OR AB toddler* | 9,472 |
| S7 | TI infant* OR AB infant* | 82,018 |
| S8 | TI ( baby or babies ) OR AB ( baby or babies ) | 16,766 |
| S9 | TI neonate* OR AB neonate* | 4,983 |
| S10 | TI ( newborn* or (new W0 born*) ) OR AB ( newborn* or (new W0 born*) ) | 11,420 |
| S11 | TI girl* OR AB girl* | 73,709 |
| S12 | TI boy* OR AB boy* | 76,589 |
| S13 | TI preschool* OR AB preschool* | 41,840 |
| S14 | TI school* OR AB school* | 391,617 |
| S15 | TI adolescen* OR AB adolescen* | 239,339 |
| S16 | TI teen* OR AB teen* | 22,205 |
| S17 | TI youth* OR AB youth* | 101,342 |
| S18 | TI juvenile* OR AB juvenile* | 25,662 |
| S19 | TI ( young W0 (person* or people) ) OR AB ( young W0 (person* or people) ) | 32,562 |
| S20 | TI kindergarten* OR AB kindergarten* | 17,935 |
| S21 | S1 OR S2 OR S3 OR S4 OR S5 OR S6 OR S7 OR S8 OR S9 OR S10 OR S11 OR S12 OR S13 OR S14 OR S15 OR S16 OR S17 OR S18 OR S19 OR S20 | 1,292,586 |
| S22 | TI ( (long or longterm) N3 (covid* or corona or coronavirus*) ) OR AB ( (long or longterm) N3 (covid* or corona or coronavirus*) ) | 30 |
| S23 | TI ( (sustain* or history) N2 (covid* or corona or coronavirus*) ) OR AB ( (sustain or history) N2 (covid* or corona or coronavirus*) ) | 12 |
| S24 | TI ( chronic N2 (covid* or corona or coronavirus*) ) OR AB ( chronic N2 (covid* or corona or coronavirus*) ) | 6 |
| S25 | TI ( prolong* N2 (covid* or corona or coronavirus*) ) OR AB ( prolong* N2 (covid* or corona or coronavirus*) ) | 189 |
| S26 | TI ( persist* N2 (covid* or corona or coronavirus*) ) OR AB ( persist* N2 (covid* or corona or coronavirus*) ) | 3 |
| S27 | TI ( (post or postacute or postvir*) N2 (covid* or corona or coronavirus*) ) OR AB ( (post or postacute or postvir*) N2 (covid* or corona or coronavirus*) ) | 40 |
| S28 | S22 OR S23 OR S24 OR S25 OR S26 OR S27 | 91 |
| S29 | DE "Coronavirus" OR DE "Severe Acute Respiratory Syndrome" | 1,863 |
| S30 | ((novel or new or nouveau) N1 (CoV or Pandemi*) | 60 |
| S31 | coronavir* or (corona W0 vir*) or OC43 or NL63 or 229E or HKU1 or HCoV* or NCoV* or covid* or (sars W0 cov*) or sarscov* | 3,228 |
| S32 | betacorona* | 411 |
| S33 | (wuhan or hubei or huanan) W0 virus | 0 |
| S34 | 2019nCoV* or CoV2 or "CoV 2" or CoV-2 | 17 |
| S35 | DE "Pneumonia" | 804 |
| S36 | wuhan or hubei or huanan | 4,539 |
| S37 | S29 OR S30 OR S31 OR S32 OR S33 OR S34 OR (S35 AND S36) | 3,480 |
| S38 | TI ((chronic or persist* or longterm or "long term" or prolong* or sustain* or continu*) N2 (complication* or infect* or symptom* or syndrome* or consequence* or outcome* or impact* or suffer* or effect* or debilit*)) OR AB ((chronic or persist* or longterm or "long term" or prolong* or sustain* or continu*) N2 (complication* or infect* or symptom* or syndrome* or consequence* or outcome* or impact* or suffer* or effect* or debilit*)) | 75,978 |
| S39 | TI ( (post or postvir* or postacute) N3 (complication* or infect* or symptom* or syndrome* or consequence* or outcome* or impact* or suffer* or effect* or debilit*) ) OR AB ( (post or postvir* or postacute) N3 (complication* or infect* or symptom* or syndrome* or consequence* or outcome* or impact* or suffer* or effect* or debilit*) ) | 14,816 |
| S40 | TI ( (long W0 haul*) or longhaul* or sequela* ) OR AB ( (long W0 haul*) or longhaul* or sequela*) | 9,988 |
| S41 | TI ((after or following) W0 infect*) OR AB ((after or following) W0 infect*) | 343 |
| S42 | TI ( (prolong* or extend* or lengthen* or postpone* or delay*) N2 (recovery or convalescen* or rehab*) ) OR AB ( (prolong* or extend* or lengthen* or postpone* or delay*) N2 (recovery or convalescen* or rehab*)) | 1,195 |
| S43 | S38 OR S39 OR S40 OR S41 OR S42 | 99,579 |
| S44 | S37 AND S43 | 211 |
| S45 | S28 OR S44 | 279 |
| S46 | S21 AND S45 | 54 |
| S47 | S21 AND S45  **Limiters** - Published Date: 20191201-20210131 | 53 |

Web of Science 05.01.2021 Web of Science (Indexes=SCI-EXPANDED, SSCI)

| # 27 | [260](https://apps-webofknowledge-com.ezproxy.keele.ac.uk/summary.do?product=WOS&doc=1&qid=28&SID=F3JCH2iV5n6HLTSIzUW&search_mode=AdvancedSearch&update_back2search_link_param=yes) | #25 AND #5  *Indexes=SCI-EXPANDED, SSCI Timespan=2019-2021* |
| --- | --- | --- |
| # 26 | [331](https://apps-webofknowledge-com.ezproxy.keele.ac.uk/summary.do?product=WOS&doc=1&qid=27&SID=F3JCH2iV5n6HLTSIzUW&search_mode=AdvancedSearch&update_back2search_link_param=yes) | #25 AND #5 |
| # 25 | [3,518](https://apps-webofknowledge-com.ezproxy.keele.ac.uk/summary.do?product=WOS&doc=1&qid=26&SID=F3JCH2iV5n6HLTSIzUW&search_mode=CombineSearches&update_back2search_link_param=yes) | #24 OR #12 |
| # 24 | [2,124](https://apps-webofknowledge-com.ezproxy.keele.ac.uk/summary.do?product=WOS&doc=1&qid=25&SID=F3JCH2iV5n6HLTSIzUW&search_mode=CombineSearches&update_back2search_link_param=yes) | #23 AND #18 |
| # 23 | [625,270](https://apps-webofknowledge-com.ezproxy.keele.ac.uk/summary.do?product=WOS&doc=1&qid=24&SID=F3JCH2iV5n6HLTSIzUW&search_mode=CombineSearches&update_back2search_link_param=yes) | #22 OR #21 OR #20 OR #19 |
| # 22 | [9,512](https://apps-webofknowledge-com.ezproxy.keele.ac.uk/summary.do?product=WOS&doc=1&qid=23&SID=F3JCH2iV5n6HLTSIzUW&search_mode=AdvancedSearch&update_back2search_link_param=yes) | ts= ((prolong* or extend* or lengthen* or postpone* or delay*) near/2 (recovery or convalescen* or rehab*) ) |
| # 21 | [53,563](https://apps-webofknowledge-com.ezproxy.keele.ac.uk/summary.do?product=WOS&doc=1&qid=22&SID=F3JCH2iV5n6HLTSIzUW&search_mode=AdvancedSearch&update_back2search_link_param=yes) | ts=("long haul*" or longhaul* or sequela* or ((after or following) W0 infect*)) |
| # 20 | [93,192](https://apps-webofknowledge-com.ezproxy.keele.ac.uk/summary.do?product=WOS&doc=1&qid=21&SID=F3JCH2iV5n6HLTSIzUW&search_mode=AdvancedSearch&update_back2search_link_param=yes) | ts=((post or postvir* or postacute) near/3 (complication* or infect* or symptom* or syndrome* or consequence* or outcome* or impact* or suffer* or effect* or debilit*) ) |
| # 19 | [482,915](https://apps-webofknowledge-com.ezproxy.keele.ac.uk/summary.do?product=WOS&doc=1&qid=20&SID=F3JCH2iV5n6HLTSIzUW&search_mode=AdvancedSearch&update_back2search_link_param=yes) | ts=((chronic or persist* or longterm or "long term" or prolong* or sustain* or continu*) near/2 (complication* or infect* or symptom* or syndrome* or consequence* or outcome* or impact* or suffer* or effect* or debilit*) ) |
| # 18 | [76,264](https://apps-webofknowledge-com.ezproxy.keele.ac.uk/summary.do?product=WOS&doc=1&qid=19&SID=F3JCH2iV5n6HLTSIzUW&search_mode=CombineSearches&update_back2search_link_param=yes) | #17 OR #16 OR #15 OR #14 OR #13 |
| # 17 | [17,688](https://apps-webofknowledge-com.ezproxy.keele.ac.uk/summary.do?product=WOS&doc=1&qid=18&SID=F3JCH2iV5n6HLTSIzUW&search_mode=AdvancedSearch&update_back2search_link_param=yes) | ts=(2019nCoV* or CoV2 or "CoV 2") |
| # 16 | [16](https://apps-webofknowledge-com.ezproxy.keele.ac.uk/summary.do?product=WOS&doc=1&qid=17&SID=F3JCH2iV5n6HLTSIzUW&search_mode=AdvancedSearch&update_back2search_link_param=yes) | ts=("wuhan virus" or "hubei virus" or "huanan virus") |
| # 15 | [2,695](https://apps-webofknowledge-com.ezproxy.keele.ac.uk/summary.do?product=WOS&doc=1&qid=15&SID=F3JCH2iV5n6HLTSIzUW&search_mode=AdvancedSearch&update_back2search_link_param=yes) | ts=((pneumonia or SARS or "severe acute respiratory") and (wuhan or hubei or huanan) ) |
| # 14 | [75,405](https://apps-webofknowledge-com.ezproxy.keele.ac.uk/summary.do?product=WOS&doc=1&qid=14&SID=F3JCH2iV5n6HLTSIzUW&search_mode=AdvancedSearch&update_back2search_link_param=yes) | ts=(coronavir* or "corona vir*" or OC43 or NL63 or 229E or HKU1 or HCoV* or NCoV* or covid* or "sars cov*" or sarscov* or betacorona*) |
| # 13 | [1,447](https://apps-webofknowledge-com.ezproxy.keele.ac.uk/summary.do?product=WOS&doc=1&qid=13&SID=F3JCH2iV5n6HLTSIzUW&search_mode=AdvancedSearch&update_back2search_link_param=yes) | ts=((novel or new or nouveau) near/1 (CoV or Pandemi*) ) |
| # 12 | [1,659](https://apps-webofknowledge-com.ezproxy.keele.ac.uk/summary.do?product=WOS&doc=1&qid=12&SID=F3JCH2iV5n6HLTSIzUW&search_mode=CombineSearches&update_back2search_link_param=yes) | #11 OR #10 OR #9 OR #8 OR #7 OR #6 |
| # 11 | [720](https://apps-webofknowledge-com.ezproxy.keele.ac.uk/summary.do?product=WOS&doc=1&qid=11&SID=F3JCH2iV5n6HLTSIzUW&search_mode=AdvancedSearch&update_back2search_link_param=yes) | ts=((post or postacute or postvir*) near/3 (covid* or corona or coronavirus*) ) |
| # 10 | [107](https://apps-webofknowledge-com.ezproxy.keele.ac.uk/summary.do?product=WOS&doc=1&qid=10&SID=F3JCH2iV5n6HLTSIzUW&search_mode=AdvancedSearch&update_back2search_link_param=yes) | ts=(persist* near/2 (covid* or corona or coronavirus*) ) |
| # 9 | [54](https://apps-webofknowledge-com.ezproxy.keele.ac.uk/summary.do?product=WOS&doc=1&qid=9&SID=F3JCH2iV5n6HLTSIzUW&search_mode=AdvancedSearch&update_back2search_link_param=yes) | ts=(prolong* near/2 (covid* or corona or coronavirus*) ) |
| # 8 | [62](https://apps-webofknowledge-com.ezproxy.keele.ac.uk/summary.do?product=WOS&doc=1&qid=8&SID=F3JCH2iV5n6HLTSIzUW&search_mode=AdvancedSearch&update_back2search_link_param=yes) | ts=(chronic near/2 (covid* or corona or coronavirus*) ) |
| # 7 | [205](https://apps-webofknowledge-com.ezproxy.keele.ac.uk/summary.do?product=WOS&doc=1&qid=7&SID=F3JCH2iV5n6HLTSIzUW&search_mode=AdvancedSearch&update_back2search_link_param=yes) | ts=((sustain* or history) near/2 (covid* or corona or coronavirus*) ) |
| # 6 | [554](https://apps-webofknowledge-com.ezproxy.keele.ac.uk/summary.do?product=WOS&doc=1&qid=6&SID=F3JCH2iV5n6HLTSIzUW&search_mode=AdvancedSearch&update_back2search_link_param=yes) | ts=(("long term" or long) near/3 (covid* or corona or coronavirus*) ) |
| # 5 | [2,976,463](https://apps-webofknowledge-com.ezproxy.keele.ac.uk/summary.do?product=WOS&doc=1&qid=5&SID=F3JCH2iV5n6HLTSIzUW&search_mode=CombineSearches&update_back2search_link_param=yes) | #4 OR #3 OR #2 OR #1 |
| # 4 | [711,073](https://apps-webofknowledge-com.ezproxy.keele.ac.uk/summary.do?product=WOS&doc=1&qid=4&SID=F3JCH2iV5n6HLTSIzUW&search_mode=AdvancedSearch&update_back2search_link_param=yes) | ts=(adolescen* or teen* or youth* or juvenile* or "young person*" or "young people*") |
| # 3 | [688,818](https://apps-webofknowledge-com.ezproxy.keele.ac.uk/summary.do?product=WOS&doc=1&qid=3&SID=F3JCH2iV5n6HLTSIzUW&search_mode=AdvancedSearch&update_back2search_link_param=yes) | ts=(girl* or boy* or school* or preschool* or kindergarten*) |
| # 2 | [260,592](https://apps-webofknowledge-com.ezproxy.keele.ac.uk/summary.do?product=WOS&doc=1&qid=2&SID=F3JCH2iV5n6HLTSIzUW&search_mode=AdvancedSearch&update_back2search_link_param=yes) | ts=(baby or babies or neonate* or newborn* or "new born*") |
| # 1 | [2,105,599](https://apps-webofknowledge-com.ezproxy.keele.ac.uk/summary.do?product=WOS&doc=1&qid=1&SID=F3JCH2iV5n6HLTSIzUW&search_mode=AdvancedSearch&update_back2search_link_param=yes) | ts=(paediatri* or pediatri* or child* or toddler* or infant*) |

ASSIA (ProQUEST) 05.01.2021 - Applied Social Sciences Index & Abstracts (ASSIA)

[noft(Paediatri* OR pediatri* OR child* OR toddler* OR infant* OR baby OR babies OR neonate* OR newborn* OR girl* OR boy* OR school* OR preschool* OR kindergarten* OR adolescen* OR teen* OR youth* OR juvenile* OR "young person" OR "young people" OR" "new born" OR "new borns") AND (noft("long covid") OR (noft(("long haul" OR "long hauler" OR "long haulers" OR sequela* OR ((prolong* OR extend* OR lengthen* OR postpone* OR delay*) AND (recovery OR convalescen* OR rehab*)) OR "long term" OR longterm OR chronic OR prolong* OR persist* OR postacute OR postvir* OR "post viral" OR "post virus" OR "post acute")) AND (noft(covid* OR corona OR coronavirus* OR OC43 OR NL63 OR 229E OR HKU1 OR HCoV* OR NCoV* OR sarscov* OR betacorona* OR 2019nCoV* OR CoV2 OR "Cov 2") OR noft("wuhan virus" OR "hubei virus" OR "hunanan virus") OR (noft(wuhan OR hubei OR hunanan) AND noft(pneumonia OR SARS OR "severe acute respiratory"))))) AND pd(20191201-20210105)](https://search.proquest.com/myresearch/savedsearches.checkdbssearchlink:rerunsearch/1862761/SavedSearches?site=assia&t:ac=SavedSearches)

Coronavirus Research Database (ProQUEST) 05.01.2021 - Applied Social Sciences Index & Abstracts (ASSIA)

[noft(Paediatri* OR pediatri* OR child* OR toddler* OR infant* OR baby OR babies OR neonate* OR newborn* OR girl* OR boy* OR school* OR preschool* OR kindergarten* OR adolescen* OR teen* OR youth* OR juvenile* OR "young person" OR "young people" OR" "new born" OR "new borns") AND (noft("long covid") OR (noft(("long haul" OR "long hauler" OR "long haulers" OR sequela* OR ((prolong* OR extend* OR lengthen* OR postpone* OR delay*) AND (recovery OR convalescen* OR rehab*)) OR "long term" OR longterm OR chronic OR prolong* OR persist* OR postacute OR postvir* OR "post viral" OR "post virus" OR "post acute")) AND (noft(covid* OR corona OR coronavirus* OR OC43 OR NL63 OR 229E OR HKU1 OR HCoV* OR NCoV* OR sarscov* OR betacorona* OR 2019nCoV* OR CoV2 OR "Cov 2") OR noft("wuhan virus" OR "hubei virus" OR "hunanan virus") OR (noft(wuhan OR hubei OR hunanan) AND noft(pneumonia OR SARS OR "severe acute respiratory"))))) AND pd(20191201-20210105)](https://search.proquest.com/myresearch/savedsearches.checkdbssearchlink:rerunsearch/1862778/SavedSearches?site=coronavirus&t:ac=SavedSearches)

Cochrane Covid-19 study register (<https://covid-19.cochrane.org/>) 05.01.2021

**Search 1**

Paediatri* or pediatri* or child* or toddler* or infant* or baby or babies or neonate* or newborn* or girl* or boy* or school* or preschool* or kindergarten* or adolescen* or teen* or youth* or juvenile* or "young person" or "young people" or "new born" or "new borns"

**AND**

"long covid" or longhaul* or "long haul" or "long hauler" Or "long haulers" or sequela*

**AND**

1 Dec '19 - 5 Jan '21

**N** = 30

**Search 2**

Paediatri* or pediatri* or child* or toddler* or infant* or baby or babies or neonate* or newborn* or girl* or boy* or school* or preschool* or kindergarten* or adolescen* or teen* or youth* or juvenile* or "young person" or "young people" or "new born" or "new borns"

**AND**

"long term" OR longterm OR chronic OR prolong* OR persist* OR postacute or postvir* or "post viral" or "post virus" or "post acute" or sustain* OR continu*

**AND**

complication* or symptom* or syndrome* or consequence* or impact* or suffer* or debilit*

**AND**

1 Dec '19 - 5 Jan '21

**N** = 674

**Search 3**

Paediatri* or pediatri* or child* or toddler* or infant* or baby or babies or neonate* or newborn* or girl* or boy* or school* or preschool* or kindergarten* or adolescen* or teen* or youth* or juvenile* or "young person" or "young people" or "new born" or "new borns"

**AND**

prolong* or extend* Or lengthen* or postpone* or delay*

**AND**

recovery or convalescen* OR rehab*

**AND**

1 Dec '19 - 5 Jan '21

**N** = 29

**Total unique** = 705

WHO Covid-19 Global Literature on coronavirus disease (<https://search.bvsalud.org/global-literature-on-novel-coronavirus-2019-ncov/>) 05.01.2021

(Paediatri* or pediatri* or child* or toddler* or infant* or baby or babies or neonate* or newborn* or girl* or boy* or school* or preschool* or kindergarten* or adolescen* or teen* or youth* or juvenile* or “young person” or “young people” or “new born” or “new borns”) AND (“long covid” or “long haul” or “long hauler” Or “long haulers” or longhaul* or sequela* or “after infection” or “following infection” Or ((prolong* or extend* Or lengthen* or postpone* or delay*) AND (recovery or convalescen* OR rehab*)) or “long term” OR longterm OR chronic OR prolong* OR persist* OR postacute or postvir* or “post viral” or “post virus” or “post acute”)

**N** = 450 all databases excluding MEDLINE (1977 including MEDLINE [n=1527])

OpenGrey (<http://www.opengrey.eu/>) 05.01.2021

coronavir* OR corona OR CoV OR wuhan OR hubei OR huanan OR OC43 OR NL63 OR 229E OR HKU1 OR HCoV* OR NCoV* OR covid* OR sarscov*

Year 2019-2021

**N** = 0

NDLTD Global ETD Search (<http://search.ndltd.org/>) 05.01.2021

(cov sars 2019n corona wuhan hubei hunanan OC43 NL63 229E HKU1 HCoV NCoV betacorona) AND (Paediatri pediatri child toddler infant baby babies neonate newborn girl boy school preschool kindergarten adolescen teen youth juvenile "young person" " young people " "new born")

**N** = 24

Supplementary material – S4 – Funnel plot

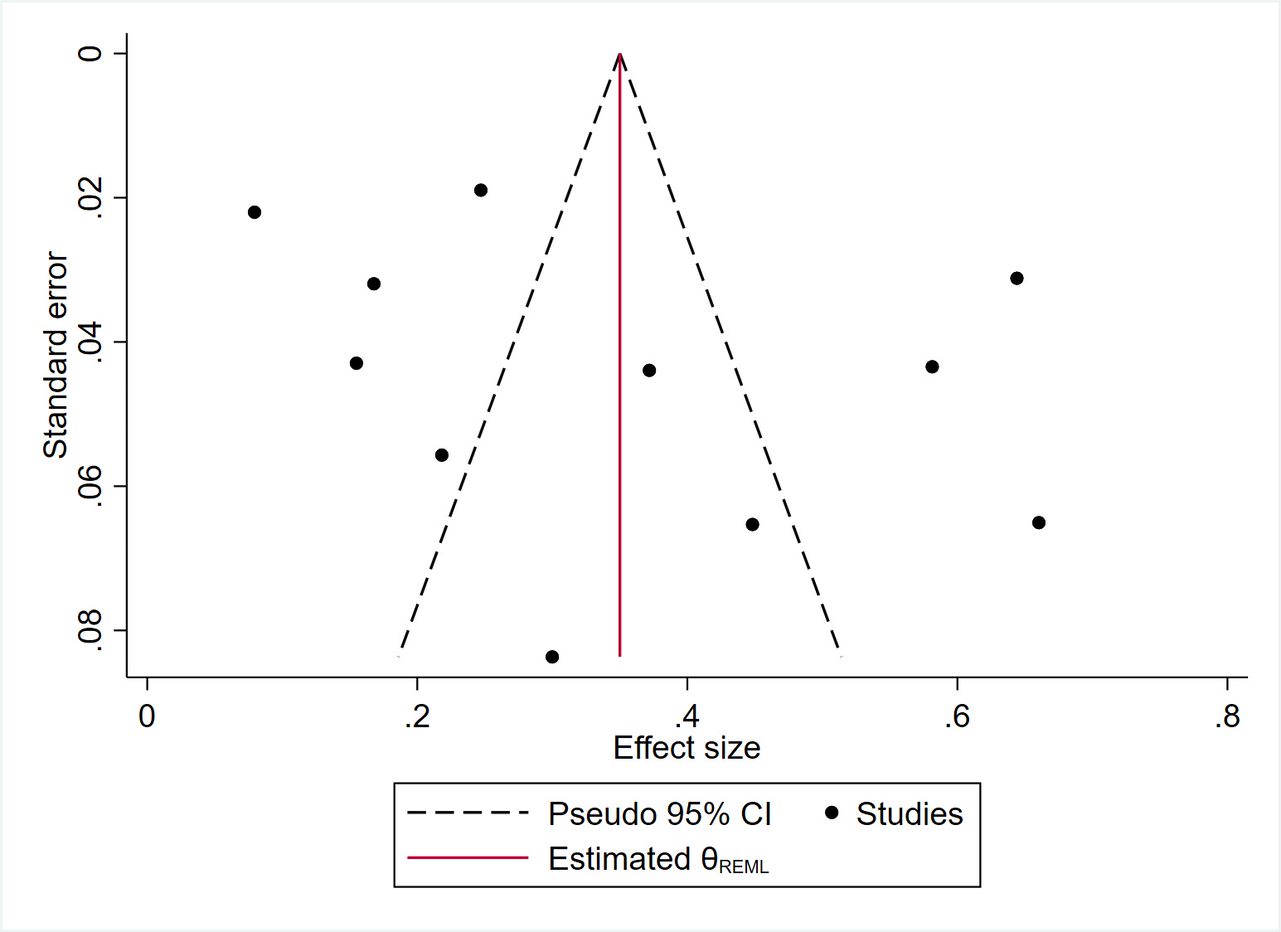
